# Host-pathogen sympatry and differential transmissibility of *Mycobacterium tuberculosis complex*

**DOI:** 10.1101/2022.08.04.22278337

**Authors:** Matthias I Gröschel, Francy J. Pérez-Llanos, Roland Diel, Roger Vargas, Vincent Escuyer, Kimberlee Musser, Lisa Trieu, Jeanne Sullivan Meissner, Jillian Knorr, Don Klinkenberg, Peter Kouw, Susanne Homolka, Wojciech Samek, Barun Mathema, Dick van Soolingen, Stefan Niemann, Shama Ahuja, Maha R Farhat

## Abstract

The obligate human pathogen *Mycobacterium* tuberculosis complex (*Mtbc*) separates genetically into nine lineages several of which demonstrate sympatry with their human host i.e. have distinct and restricted patterns of geographical distribution globally.^1–3^ Geographically restricted *Mtbc* lineages have been hypothesized to be adapted to infect and/or transmit among sympatric human hosts, *i*.*e*. to be niche specialists, but this is yet to be confirmed while controlling for exposure, social networks and risk of disease after exposure.^1,4^ Here we show that strains of geographically restricted (*Mtbc* lineages L1,L2_restricted_, L3,L4_restricted_, L5,L6 are intrinsically less transmissible than widespread *Mtbc* lineages (L2_widespread_, L4_widespread_) across Western European and North American cosmopolitan populations. Comparing transmissibility between sympatric and allopatric contact-pathogen pairs, we find the first controlled evidence for a biological impact of sympatry between *Mtbc* strains and their human hosts; allopatric host-pathogen exposures has a 38% decrease in the odds of infection among contacts compared with sympatric exposures. We measure 10- fold lower phagocytosis and growth rates of L6 geographically restricted strains compared to L4_widespread_ in *in vitro* allopatric macrophage infections. Long-term co-existence of *Mtbc* strains and humans has resulted in differential transmissibility between allopatric and sympatric hosts for strains of geographically restricted lineages. Understanding the specific genetic and immunological underpinnings of sympatry in TB may inform rational vaccine design and TB control.

## INTRODUCTION

Humans and their ancestors have continuously inhabited an earth replete with a wide diversity of microbial species. While many of these species are harmless or even beneficial to humans, some have the ability to infect human tissues and inflict harm, including pathogenic species that evolved the concurrent ability to transmit between humans (e.g., via contact, droplets or airborne transmission). Until recently, the study of host-pathogen relationships at long-time scales was methodologically limited to model organisms in model environments, as direct investigation of naturally occurring pathogen adaptation and long- term generational effects was technically and logistically difficult.^5^

High-throughput whole genome sequencing (WGS) has since enabled the direct study of hundreds of thousands of pathogens sampled directly from their human host, including pathogens of the *Mycobacterium tuberculosis* complex (*Mtbc*). Strains of the genetic lineages of the *Mtbc* exhibit distinct geographical distributions across the world regions:^6^ Tuberculosis (TB) disease in Europe and North-America is mainly caused by Lineage (L) 4 (Euro-American) strains and specifically the three sublineages 4.1.2/Haarlem, 4.10/PGG3 and L4.3/LAM.^2^ *Mtbc* sublineage 4.1.1/X has mostly been isolated from the Americas and sublineages 4.1.3/Ghana, 4.6.1/Uganda and 4.6.2/Cameroon are largely restricted to sub- Saharan Africa.^1–3^ TB disease in Asia is usually caused by the Beijing lineage (L2, more than 70% in China). ^3,7^. In West-Africa, TB disease can be caused by *M. africanum* West African 1 and 2 (*Mtbc* Lin5 and 6), lineages phylogenetically ancestral to *Mtb* sensu stricto (Lineages 1-4 & L7). *Mtbc* L5 and L6 have to-date not been isolated outside of West-Africa.^8,9^ Similarly, *Mtbc* L7 and 8 have only been isolated in East Africa.^10,11^

It has been suggested that as an obligate human pathogen, Mtbc, co-diverged with its human host and the resulting geographical spread of its genetic lineages is a result of co- evolution.^1,12^ However, this would place bacterial evolution on a similar timescale as that of humans. This possibility has been questioned in other works including those that applied formal tests for co-divergence to small contemporaneous samples of Mtb and representative human Y chromosome data.^13–17^ With or without co-divergence, studies describing the phylo- geographical distribution of *Mtbc*, strains within geographically-restricted lineages of *Mtbc* are hypothesized to preferentially infect local human hosts with a restricted range of genotypes, while strains of other lineages are isolated worldwide, capable of infecting hosts with a wide range of genotypes.^1–3,6,18–20^ The geographic overlap of two distinct species is also termed sympatry, but it has not been studied whether the long term co-existence of *Mtbc* lineages and local hosts has resulted in any biological effect that could explain the geographic restriction and global spread of the *Mtbc* lineages.

Intriguingly, stable pathogen-host ancestry associations among secondary TB cases have also been observed in two cosmopolitan cities where mixing of host and pathogen populations occurs, suggesting different transmission dynamics attributable to both pathogen and host ancestry.^4,21^ However, as *Mtbc* is an airborne pathogen, observations suggesting a biological effect resulting from long term co-existence of host and pathogen have been alternatively attributed to differential exposure to *Mtbc* lineages as a result of social mixing patterns, genetic drift, or founder effects rather than an effect of host-pathogen sympatry.^1^ These studies often measured transmission using data derived only from individuals with TB after the development of active disease and did not control for social mixing or other aspects of exposure.

Here, we set out to study if the observed sympatry (geographic co-localization) between *Mtbc* and local hosts has a measurable impact on the TB epidemic. To test the hypothesis whether geographically restricted lineages can infect hosts with a restricted range of genotypes based on geographic co-localization, while widespread lineages can infect hosts with a wider range of genotypes, we leverage detailed public health contact tracing data and whole genome sequencing from three low TB incidence cosmopolitan cities. We first study transmissibility between geographically restricted and globally spread *Mtbc* strains while controlling for founder effects, host factors, and extent of exposure. We then quantify the effect of sympatry on transmissibility, and follow up the epidemiological analysis with *in vitro* macrophage infection experiments.

## RESULTS

### A multi-site cosmopolitan cohort of tuberculosis cases and close contacts

A major barrier to investigating a biological role of sympatry in *Mtbc* is the limitation in size and depth of contact tracing data directly measuring transmissibility in cosmopolitan societies with mixing of human and pathogen populations. To address this, we built a large cohort of TB cases (n = 5,256) and their contacts (n=28,889) from New York City (NYC), Amsterdam, and Hamburg. The three settings routinely and systematically surveyed for TB cases and contacts as part of TB control activities and performed whole genome sequencing (WGS) for strains from nearly all active culture-positive TB cases in this cohort (>99%, 5,219/5,256). WGS allowed *Mtbc* lineage classification and informed transmission assessment. In the study of transmissibility, we were able to control for founder effects by considering close contacts of each active TB index case individually. We were able to control for exposure by modeling transmissibility using variables of TB index case infectiousness and contact exposure time.

After excluding extrapulmonary TB as potential source cases, TB index cases without linked contacts, and removing contacts without significant exposure (see **Methods** for exposure times considered significant by site), the transmission analysis cohort comprised 2,279 patients with pulmonary TB linked to 12,749 close contacts (**Methods, Table 1, Figure 1a**). As extrapulmonary TB cases without clinical or bacterial evidence of pulmonary TB are not thought to be contagious they were excluded as potentially transmitting index cases. The median number of close contacts investigated per index TB case was respectively 4 contacts (IQR 2 – 6), 4 (IQR 2 – 8), and 3 (IQR 1 – 7) in NYC, Amsterdam, and Hamburg (**Figure 1b**).

**Table 1:** Pulmonary tuberculosis patients characteristics.

|  | N | Overall<br>N = 2,279<sup>1</sup> | Hamburg<br>N = 498<sup>1</sup> | Netherlands<br>N = 266<sup>1</sup> | New York City<br>N = 1,515<sup>1</sup> |
| --- | --- | --- | --- | --- | --- |
| **Sex** | 2,279 |  |  |  |  |
| Female |  | 849 (37%) | 191 (38%) | 98 (37%) | 560 (37%) |
| Male |  | 1,430 (63%) | 307 (62%) | 168 (63%) | 955 (63%) |
| **Age** | 2,279 | 48 (31, 66) | 43 (29, 61) | 38 (26, 55) | 52 (33, 68) |
| **Country of birth** | 2,279 |  |  |  |  |
| High TB incidence |  | 1,574 (69%) | 224 (45%) | 158 (59%) | 1,192 (79%) |
| Medium/low TB incidence |  | 705 (31%) | 274 (55%) | 108 (41%) | 323 (21%) |
| **Sputum microscopy** | 2,279 |  |  |  |  |
| Negative |  | 809 (35%) | 230 (46%) | 90 (34%) | 489 (32%) |
| Positive |  | 1,470 (65%) | 268 (54%) | 176 (66%) | 1,026 (68%) |
| **Symptomatic** | 2,254 | 1,985 (88%) | 448 (92%) | 203 (81%) | 1,334 (88%) |
| **Homeless** | 2,275 | 72 (3.2%) | 32 (6.5%) | 7 (2.6%) | 33 (2.2%) |
| **Drug use** | 2,279 | 108 (4.7%) | 13 (2.6%) | 6 (2.3%) | 89 (5.9%) |
| **Alcohol use** | 2,272 | 137 (6.0%) | 77 (16%) | 5 (1.9%) | 55 (3.6%) |
| **HIV** | 1,975 | 77 (3.9%) | 21 (4.7%) | 8 (3.6%) | 48 (3.7%) |
| **Diabetes** | 2,279 | 418 (18%) | 24 (4.8%) | 22 (8.3%) | 372 (25%) |
| **Immunosuppression** | 1,829 | 44 (2.4%) | 6 (3.4%) | 6 (4.3%) | 32 (2.1%) |
| **Isoniazid monoresistance** | 2,278 | 242 (11%) | 35 (7.0%) | 25 (9.4%) | 182 (12%) |
| **Multidrug resistance** | 2,278 | 60 (2.6%) | 13 (2.6%) | 3 (1.1%) | 44 (2.9%) |
| ***M. tuberculosis* complex lineage** | 2,279 |  |  |  |  |
| Geographically widespread |  | 1,636 (72%) | 396 (80%) | 183 (69%) | 1,057 (70%) |
| Geographically restricted |  | 643 (28%) | 102 (20%) | 83 (31%) | 458 (30%) |

<sup>1</sup>n (%); Median (IQR); High TB incidence defined as >30/100.000 incidence based on 2016 WHO data

**Figure 1:**
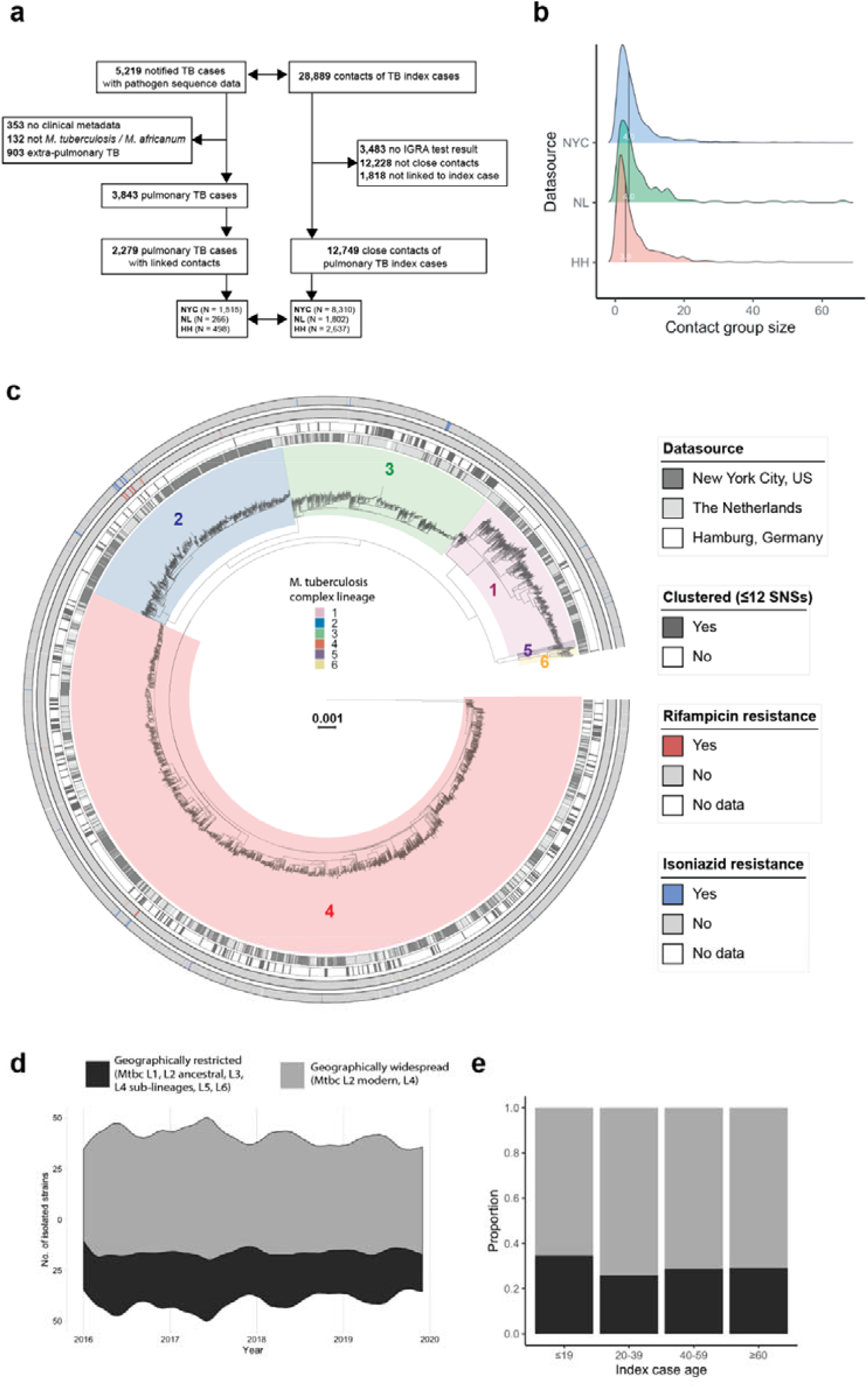
Properties of circulating *M. tuberculosis* complex geographically restricted and widespread strains. a) Flowchart of included tuberculosis index cases and linked contacts; b) Density plot of contact group sizes per city; c) Maximum likelihood phylogenetic tree of included strains, clades are coloured by genetic lineage, the rings denote (from inner to outer) the city, clustering based on a 12 Single Nucleotide Substitution cut-off, phenotypic drug resistance to Rifampicin and Isoniazid; d) Proportion of isolated strains by geographically restricted or widespread *Mtbc* lineages over the study period; e) Bar plot showing the proportions of isolated restricted or widespread *Mtbc* lineages by age groups. TB = tuberculosis, IGRA = Interferon Gamma Release Assay, SNS = Single Nucleotide Substitution, Mtbc = *M. tuberculosis* complex.

### Patient characteristics differed across study sites

The three source cities differed in key drivers of TB case burden and transmission, including birth in a country with high TB incidence, proportion of TB in the adolescent to young adult age group, sputum *Mtbc* smear positivity, alcohol or substance use disorder, and homelessness (**Table 1**).^22^ On average across study sites, 69% of TB cases and 48% of close contacts were born in a country with a high TB incidence country, defined as >30/100,000 TB incidence based WHO’s 2016 data;^23^ TB patients and close contacts from NYC were more likely to be born in a high-incidence country (Chi squared, p<0.001 for both comparisons corrected for the false discovery rate). TB patients in NYC were older by 9 and 14 years on median compared with patients in Hamburg or Amsterdam, respectively (Wilcoxon rank sum, p<0.001). TB patients from Hamburg were less often sputum smear positive (Chi squared, p=0.002 for Amsterdam and p<0.001 NYC), but more often used alcohol (p < 0.001 for both comparisons), or were homeless (p = 0.036 Amsterdam, p < 0.001 NYC). NYC TB cases more often used illicit substances (p = 0.009 Hamburg, p = 0.018 Amsterdam). Factors recognized to increase the contact risk of progression to active TB after infection, including HIV and other immunosuppression, did not differ by city. Diabetes was more common among TB cases in NYC (p < 0.001 for both comparisons). TB patients in NYC were more often isoniazid mono-resistant than in Hamburg (p = 0.005); there was no difference in multidrug resistance (MDR, resistance to isoniazid and rifampicin) across the cities.

### Geographically restricted lineages cause a measurable proportion of TB disease burden

We next used the WGS data to characterize the circulating *Mtbc* lineages in the three cities. Among the 5,087 isolate sequences from patients infected with *Mtbc sensu stricto* or *M. africanum* (including extrapulmonary TB cases and those without linked contact information), we identified 297,061 single nucleotide substitutions (SNSs) that allowed the calculation of a maximum likelihood phylogeny (**Figure 1c**). Using canonical signature SNSs, the *Mtbc* strains analyzed were classified into six of the nine known human-adapted *Mtbc* complex lineages:^18^ the most common being strains of the Euro-American Lineage (L4, n = 2,976, 59%), followed in order by strains of the East Asia Lineage (L2, n = 802, 16%), East Africa Central Asia (L3, n = 709, 14%), and East African, The Philippines, rim of the Indian Ocean lineages (L1, n = 551, 11%). A minority of strains belonged to *M. africanum* 1 (L5, n = 17) or 2 (L6, n = 32). We categorized *Mtbc* strains as geographically restricted either if their Simpson’s diversity index was ≤0.3 (ancestral branches of L2: 2.1 ‘Proto-Beijing’, 2.1.2.2.1 ‘Asia Ancestral 2’, and L4 sub-lineages: 4.11, 4.2.1.1, 4.3.i2, 4.5, 4.6.2.2 ‘Cameroon’, 4.6.1.1.1 ‘Uganda’)^18^ or if they belonged to lineages found to be phylo-geographically restricted in previous literature^1,12,18–20^ (L1, L3, L5, L6 as well as the ancestral L2 branch 2.2.2 ‘Asia Ancestral 1’) (**Suppl. Table S1**) and compared them to the remaining widespread sublineages of L2 and L4.^1,2,18^

Of 2,279 pulmonary TB patients, 28% (n = 643) were infected with strains of a geographically restricted lineage. The proportion of restricted strain infections remained stable over the study period, and there was no association with patient age or sex (Chi squared, p>0.17, **Figure 1d-e**). Strains of geographically restricted lineages were equally likely to harbor isoniazid mono-resistance (p = 0.8) and slightly less likely to harbor MDR compared with widespread strains (OR 0.98, CI_95_ 0.97 to 1, Wald p = 0.021), but the overall rate of MDR was low (n = 60, 2.6%, **Figure 1c**).

### Strains of geographically restricted *Mtbc* lineages differ in phylogenetic metrics of transmissibility

We hypothesized that strains of geographically restricted lineages were less transmissible than widespread strains in the cosmopolitan source communities. To investigate this, we compared genetic diversity metrics between strains of geographically restricted and widespread lineages using the total sample phylogeny. Terminal branch lengths, which represent an upper bound of evolutionary time since transmission for each *Mtbc* case, were significantly longer for strains of geographically restricted lineages (median 66 SNSs, Interquartile Range (IQR) 22 to 111, vs 41 SNSs, IQR 10 to 76) for widespread lineages, p < 2.2 × 10^−16^ using Wilcoxon rank sum test, **Figure 2a**). This difference was driven by a combination of the long terminal branch lengths of strains of L1 (median 80 SNSs, IQR 52 to 129), L2_restricted_ (91 SNSs, IQR 50 to 133) and L4_restricted_ (68 SNSs, IQR 22 to 109) compared to shorter branch lengths for L2_widespread_, L3, and L4_widespread_ (medians 52 (IQR 26 to 78), 38 (IQR 6 to 91), 36 (IQR 7 to 74) SNSs, respectively) (**Suppl. Figure S1a**). Strains of L2_restricted_ and L4_restricted_ had longer terminal branch lengths than strains of L2_widespread_ and L4_widespread._ Respectively (p < 3.9x 10^−8^ Wilcoxon rank sum test for both comparisons). Possible- transmission clusters were defined as strains within 12-SNSs distance of each other based on an updated filtering scheme that allows to call genetic variation across ∼7% more positions in the *Mtbc* genome compared to previous filtering scheme (**Methods**).^24^ Clustering was more common for strains of geographically widespread lineages than restricted lineages (822/3,778 of widespread lineage strains clustered in 239/292 total clusters vs 240/1,309 of restricted lineage strains clustered in 53/292 clusters, p = 0.035, Chi Squared, **Figure 2b**).

**Figure 2:**
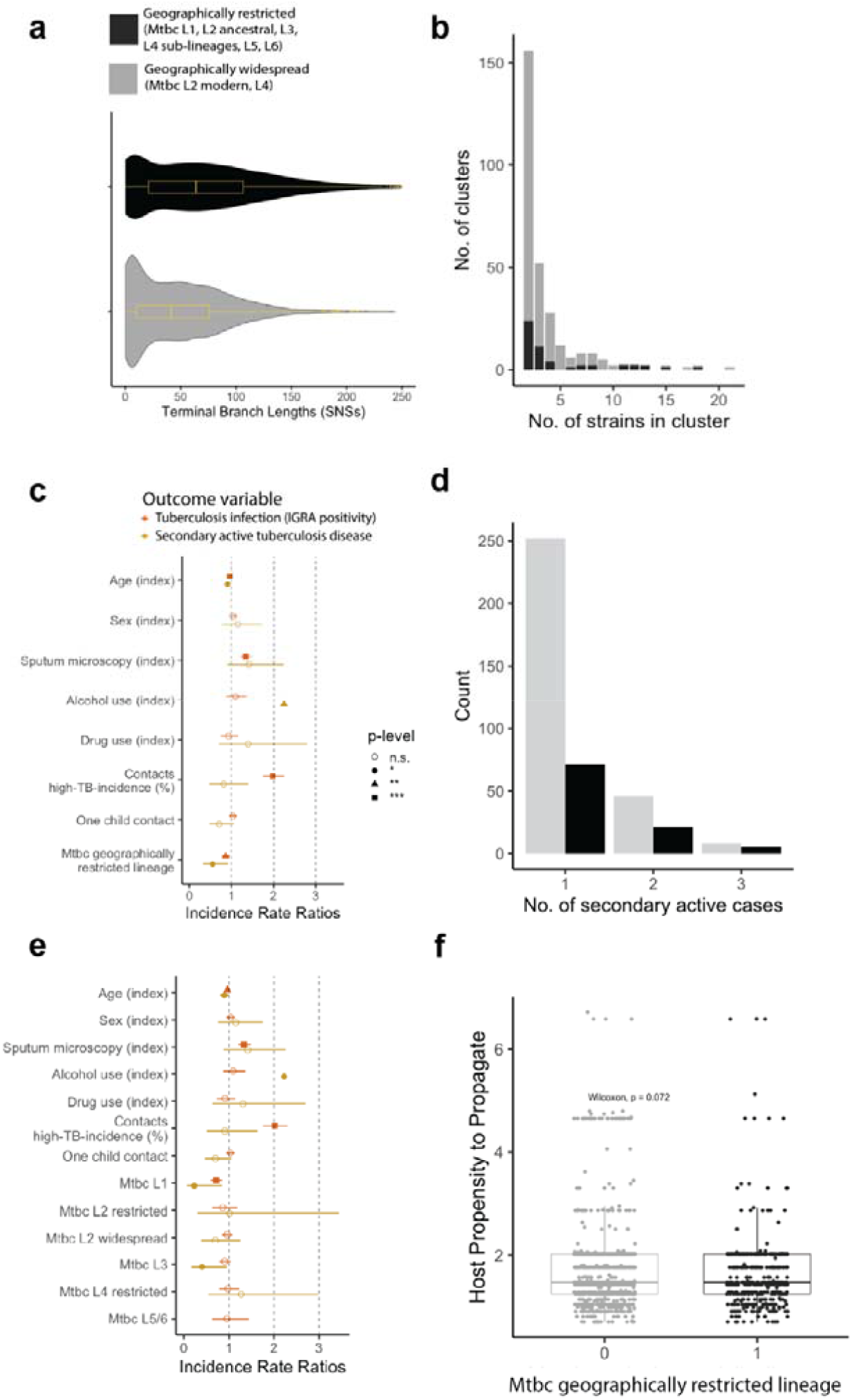
*M. tuberculosis* complex strain and tuberculosis index case characteristics associated with transmissibility and clustering. a) Violin plot of the terminal branch lengths of *Mtbc* strains, median and interquartile ranges are represented by overlaying box plots; b) Numbers of genetic clusters and number of clustered strains by geographically restricted or widespread *Mtbc* lineage (legend as in a); c) Forest plot of a multivariable Poisson rate model quantifying the effect of several predictor variables (y-axis) on the outcome ‘Count of tuberculosis infected contacts’ (in orange) or ‘Count of secondary active tuberculosis cases among contacts’ (in yellow); d) Bar plot of secondary active tuberculosis among close contacts inferred by Phybreak stratified by geographically restricted or widespread *Mtbc* lineage (legend as in a)); e) Forest plot of a multivariable Poisson rate model quantifying the effect of Mtbc lineage on tuberculosis infection (orange) or secondary active tuberculosis (yellow) among close contacts (same legend as in c); f) Host Propensity to Propagate by geographically restricted or widespread Mtbc lineage; f) Host propensity to propagate stratified by *M. tuberculosis* complex lineage. Mtbc = *M. tuberculosis* complex

The geographically restricted lineages L5/L6 comprised one L6 12-SNS cluster with two strains. The average size of clusters did not significantly differ between geographically widespread lineages compared with restricted lineages (3.4 patients vs 4.5 patients per cluster, p = 0.07, two-sample t-test, and in a sensitivity analysis using a 5-SNSs distance 3.5 patients vs 4.6 patients per cluster, p = 0.08).

### Strains of geographically restricted *Mtbc* lineages demonstrate intrinsically lower transmissibility

We isolated the effect of the infecting *Mtbc* lineage on the rate of infection among close contacts from other exposure characteristics and contact susceptibility as follows: we focused on TB infection as measured by a positive QuantiFERON®-TB Gold In-Tube test (IGRA ≥0.35 IU/ml) among close contacts (**Methods**), and (1) modeled the count of new TB infections in each index cases’ group of close contacts as a quasi-Poisson distributed random variable with the mean estimated as a linear function of *Mtbc* lineage and index case characteristics and a subset of contact susceptibility characteristics that was available across the three source sites, and (2) modelled the probability of TB infection of each individual close contact as a function of index case infectiousness and a subset of contact susceptibility variables using generalized estimating equation (GEE) with a logistic link function (**Methods**). The latter approach allowed for more fine-grain control of contact characteristics but is expected to have lower power than the quasi-Poisson model. Exposure to a geographically restricted lineage strain was associated with a lower rate of TB infections among close contacts (adjusted incidence rate ratio (IRR_adj_) 0.86, CI_95_ 0.78 to 0.95, Wald P-value = 0.004) (**Table 2, Figure 2c**). We adjusted for a higher proportion of close contacts born in a high TB-incidence country as a confounder and found its effect on IGRA positivity to be significant (IRR_adj_ 1.98, CI_95_ 1.75 to 2.25, Wald P-value < 0.001). Host predictors of TB infection among close contacts that were significantly associated in the model included younger age and sputum smear positivity of the index case (**Table 2, Figure 2c**). In the logistic GEE, exposure to a geographically restricted lineage was associated with a lower odds of TB infection among close contacts (OR_adj_ 0.82, CI_95_ 0.72 to 0.94, Wald P-value = 0.004), in agreement with the models above (**Suppl. Table S2**).

**Table 2:**
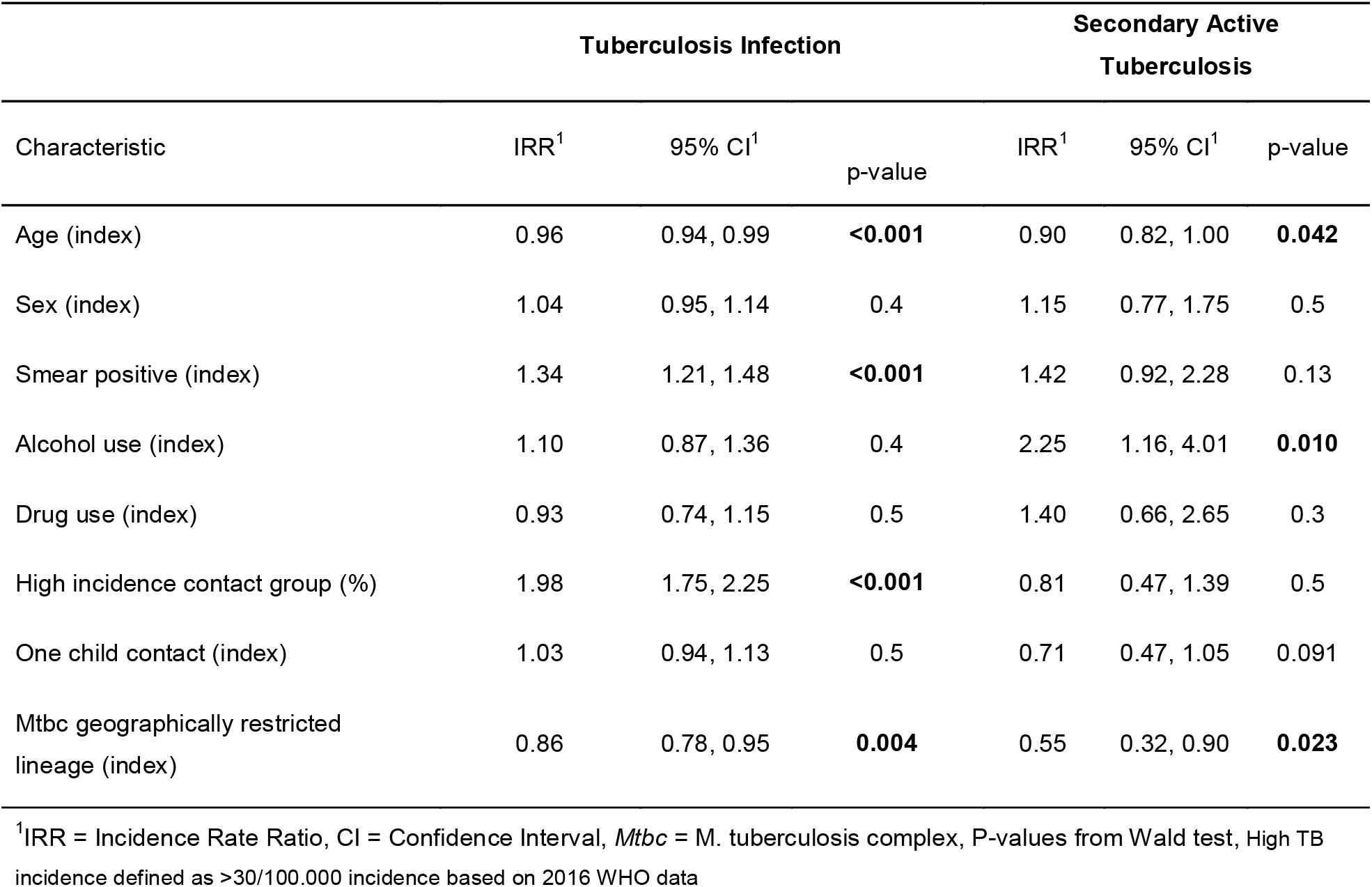
Associations of participant characteristics and bacterial factors on transmission. (quasi-Poisson model with overdispersion φ = 1.2 and φ = 1.4, respectively, N = 2,279)

We similarly modeled the interplay of *Mtbc* lineage and host characteristics on the rate of secondary active TB among close contacts using a quasi-Poisson model. Secondary pulmonary and extrapulmonary cases were linked to the source case based on a Bayesian posterior probability of transmission ≥0.5 estimated using Phybreak that uses genetic distance, sampling interval, and priors on incubation period and within-host pathogen dynamics (**Methods**).^25^ Phybreak estimated that 364 of the 2,279 pulmonary TB index cases had one close contact progress to active disease, 53 had two close contacts progress, and 10 had three progress (**Figure 2d**). Most of these transmitting TB index cases (77%, 322/427) were infected with a geographically widespread *Mtbc* lineage strain. Exposure to a geographically restricted lineage was associated with significantly lower risk of secondary active TB cases among close contacts controlling for index characteristics (IRR_adj_ 0.55, CI_95_ 0.32 to 0.90, Wald P-value = 0.023) (**Table 2, Figure 2c**). In the logistic GEE, the exposure to a geographically restricted lineage had a similar trend towards lower risk of secondary active TB but was not statistically significant (OR_adj_ 0.77, CI_95_ 0.57 to 1.04, Wald P-value = 0.09) (**Suppl. Table S2**). We used the effect estimates of all variables of the quasi-Poisson model of disease progression to calculate a host infectiousness score.^22^ The infectiousness score did not differ between index cases infected with strains of geographically restricted or widespread *Mtbc* lineages (Wilcoxon signed-rank test, p = 0.072), supporting the conclusion that host factors alone do not explain the observed differences in transmissibility (**Figure 2f**).

The above models only partially account for the effect of contact susceptibility on TB transmission and disease progression, as data on contact characteristics was limited for contacts in Hamburg and Amsterdam. We hence performed a secondary logistic GEE analysis using only NYC data to confirm the association between geographically restricted *Mtbc* lineages and intrinsic transmissibility controlling for the additional characteristics of diabetes and HIV (n = 8,481 close contacts). The model confirmed young age, male sex, and diabetes as independent predictors of secondary active TB among close contacts (**Suppl. Table S3**). HIV status among contacts was not associated with secondary active TB possibly because the majority of contacts had well controlled HIV (78% of all estimated people with HIV in New York City were virally suppressed based on data from 2020).^26^ Only four contacts reported using illicit drugs and hence this variable was not included in the model. Exposure to a geographically restricted *Mtbc* lineage was associated with lower odds of secondary active TB than estimated using the logistic GEE on the larger cohort with control for a subset of contact characteristics (OR_adj_ 0.54, CI_95_ 0.37 to 0.78, Wald P-value = 0.001 *vs*. OR_adj_ 0.77, CI_95_ 0.57 to 1.04, Wald P-value = 0.09). The effect estimate did not reach statistical significance for the model predicting TB infection (**Suppl. Table S3**).

We next evaluated the specific contributions of *Mtbc* sub-lineages on the observed differential transmissibility between geographically restricted and widespread strains across the three cosmopolitan cities using the quasi-Poisson model. L1 strains were associated with a lower rate of TB infections and secondary active cases among close contacts compared to L4_widespread_ strains (IRR_adj_ 0.71, CI_95_ 0.60 to 0.85, P-value < 0.001, and 0.23, CI_95_ 0.05 to 0.69, P-value = 0.027 respective) (**Figure 2e, Suppl. Table S4**). Although L3 strains demonstrated high proportions of clustering based on genetic distance in the Netherlands (**Suppl. Figure S1b**), the adjusted incidence rate ratio of L3 strains was 0.41 (CI_95_ 0.16 to 0.88) indicating a lower rate of secondary active cases among close contacts compared with L4_widespread_ strains (P-value < 0.032, **Figure 2e, Suppl. Table S4**). We did not include close contacts exposed to L5/L6 strains in models of secondary active TB due to the small numbers with the outcome (one of the 136 close contacts developed secondary active TB).

### Allopatric host-pathogen relationships decrease susceptibility of TB infection

The observation of lower transmissibility of strains of geographically restricted lineages in Western cosmopolitan cities may relate to their poor ability to transmit among allopatric human populations, i.e., where there has not been long-term host-pathogen co-localization. To investigate this, we first mapped countries linked to geographically restricted *Mtbc* lineages and assessed previously reported stable host ancestry-*Mtbc* lineage combinations among the TB index cases of the present analysis (**Figure 3a-b**).^4,21^ Seventy-five percent of index cases were infected with a sympatric *Mtbc* geographically restricted strain, significantly more than expected by chance (**Figure 3c-d**). To add validity to our definition of sympatry, we evaluated the geographic origin and lineage of 25,243 *Mtbc* strains with publicly available WGS (**Suppl. Figure S2a)**. To test this co-localization hypothesis, we measured the transmissibility of geographically restricted *Mtbc* lineages among exposed sympatric *vs*. allopatric close contacts using contact place of birth as a proxy for host ancestry (**Methods**).

**Figure 3:**
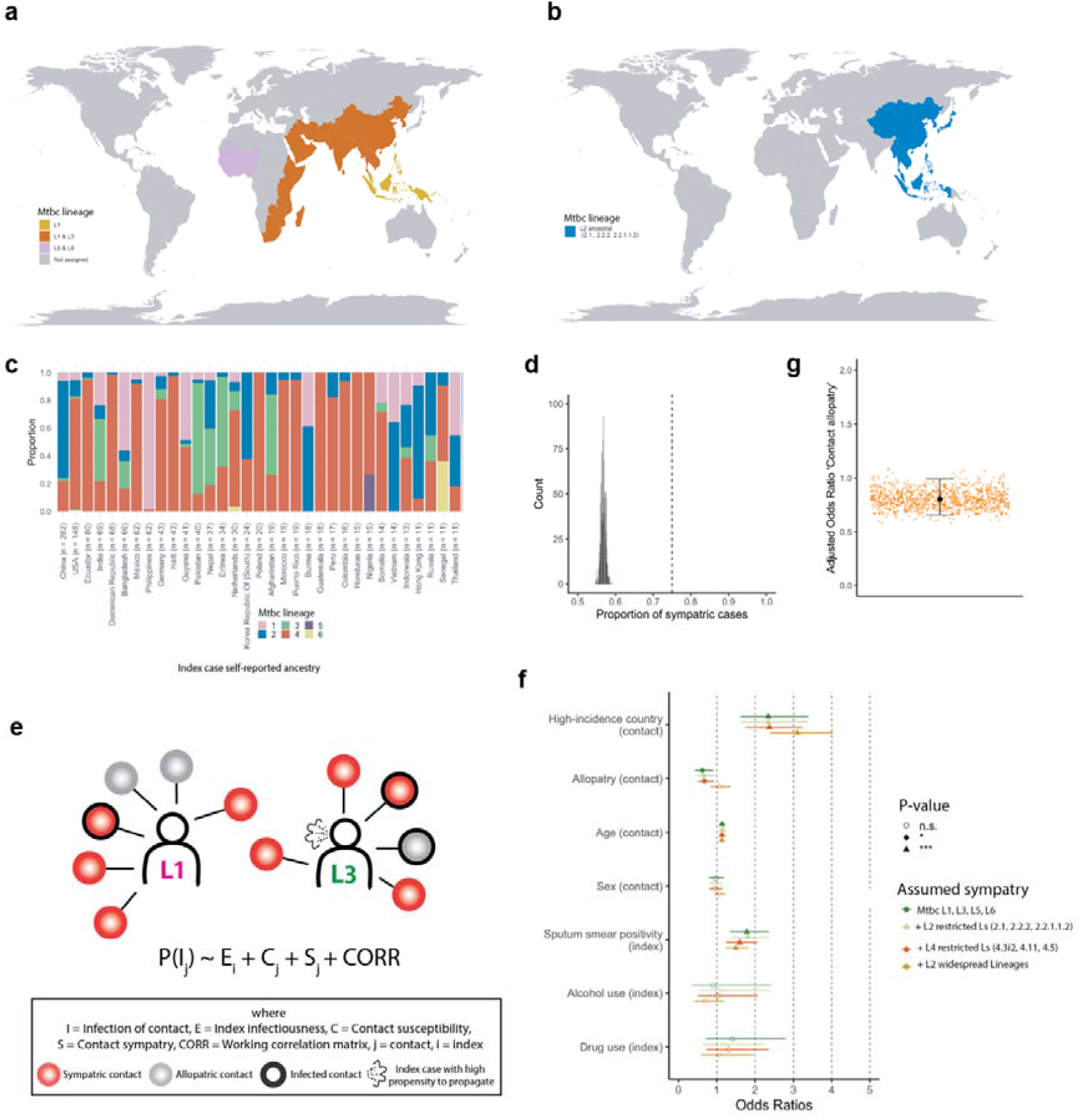
*M. tuberculosis* complex sympatry and its effect on transmissibility. a - b) Global maps displaying the countries that were considered endemic (or sympatric) to the geographically restricted *Mtbc* lineages detailed in the figure legends. For maps of the distribution of restricted Lineage 4 sublineages see Reference ^18^. c) Bar plot showing the proportions of *Mtbc* lineages isolated from the 2,279 pulmonary tuberculosis index cases included in this study shown for index cases with counts > 10; d) Permutation analysis (n = 1000) of the expected proportion of sympatric host ancestry – *Mtbc* lineage proportions. Vertical dashed line represents the proportion (0.75) among the index cases included in the study; e) Modelling approach to quantify the probability of infection for sympatric and allopatric contact exposures while including index case characteristics of infectiousness, contact characteristics of susceptibility and the correlation introduced by contact tracing. f) Forest plot of the effects estimated using a Generalized Estimating Equation model to quantify the effect of host country of birth - pathogen lineage co- localization (sympatry or allopatry). The outcome is tuberculosis infection per contact, and sympatry was defined using the maps in panel a) and b). Four models assuming four different co-localized lineage-host country of birth combinations are presented by different colors. High-incidence country defined as >30/100.000 tuberculosis incidence based on WHO 2016 data. g) Permutation test assuming 14% mixed ancestry in the sample (flipping the sympatry variable from 1 to 0 and vice versa in 14% of the sample) while rerunning the model in panel f). Mtbc = Mycobacterium tuberculosis complex.

We used a logistic GEE to study the effect of contact sympatry on infection with a geographically restricted *Mtbc* strain controlling for index and other contact characteristics (**Figure 3e**). We excluded 108/12,749 close contacts with mixed ancestry, and 667/12,749 close contacts without information on country of birth, and excluded contacts exposed to a geographically widespread lineage (L2_widespread_ and L4_widespread_). Exposure location (e.g. household vs work) did not differ between allopatric and sympatric exposures (**Suppl. Table S5**). Contact allopatry was associated with a lower odds of *Mtbc* strain infection among close contacts of index cases infected with a geographically restricted lineage strain (OR_adj_ for pooled L1,L3, L5,L6: 0.62, CI_95_ 0.43 – 0.91, n = 2556, p = 0.013, N by lineage: L1=1,080, L3=1,353, L5=35, L6=88, **Figure 3f, Suppl. Figure S2b, Suppl. Table S6**) with similar additional host factors associated with *Mtbc* strain infection as measured above in models comparing geographically restricted strains across all contacts. The effect of contact allopatry was stable when adding contacts exposed to L2_restricted_ or L4_restricted_, but extinguished when assuming the modern widespread L2 sub lineages to be sympatric to East Asian ancestry, in line with evidence of modern L2’s global spread (Suppl. Table S6).^27^ We found the association of allopatry to *Mtbc* strain infection to be robust to ancestry misclassification due to e.g. second-generation immigration in a sensitivity analysis (OR_adj_ 0.8, CI_95_ 0.65 – 0.99) (**Methods, Figure 3g**) including among the subset of data from NYC with the additional contact susceptibility variables diabetes, and HIV (**Suppl. Table S7**).

### Human macrophages differ in susceptibility to infection with *Mtbc* L4 and *M. africanum* strains by self-reported ancestry

We looked for evidence of a biological effect of sympatry using *in vitro* macrophage infection models using strains of different *Mtbc* lineages. Human blood monocyte derived macrophages (MDMs) were obtained from donors of self-identified German, Nigerian, Cameroonian or Ghanaian ancestry (see **Figure 4a** for the approach). We compared the *Mtbc* phagocytosis/uptake of two clinical isolates of the geographically widespread L4 LAM lineage with that of two clinical strains of the geographically restricted *Mtbc* L6 lineage while including the ATCC reference strains of each lineage in each group (**Suppl. Figure S3**). We ensured equal infection doses by comparing colony forming units (CFUs) of the inoculum. Macrophages of donors of self-reported ancestry co-localizing with L6 phagocytosed strains of L6 on average at 30.4% compared to 2.9% among macrophages of donors whose self-reported ancestry was Germany, not co-localizing with L6 (p < 0.001, unpaired T-test) (**Figure 4b**). There was no significant difference in growth of L4 strains in macrophages of donors of ancestry that do or do not co-localize with L6 (33.9% vs 29.5%, p = 0.8) (**Figure 4c**). This difference in uptake of L6 strains between L6 co-localizing and non-co-localizing donors was driven by all three L6 lineages (all comparisons p < 0.001) (**Figure 4d**) while we found both intra host and inter host variability in uptake efficiency of the three L4 and L6 strains (**Figure 4e, Suppl. Figure S4 and S5**).

**Figure 4.**
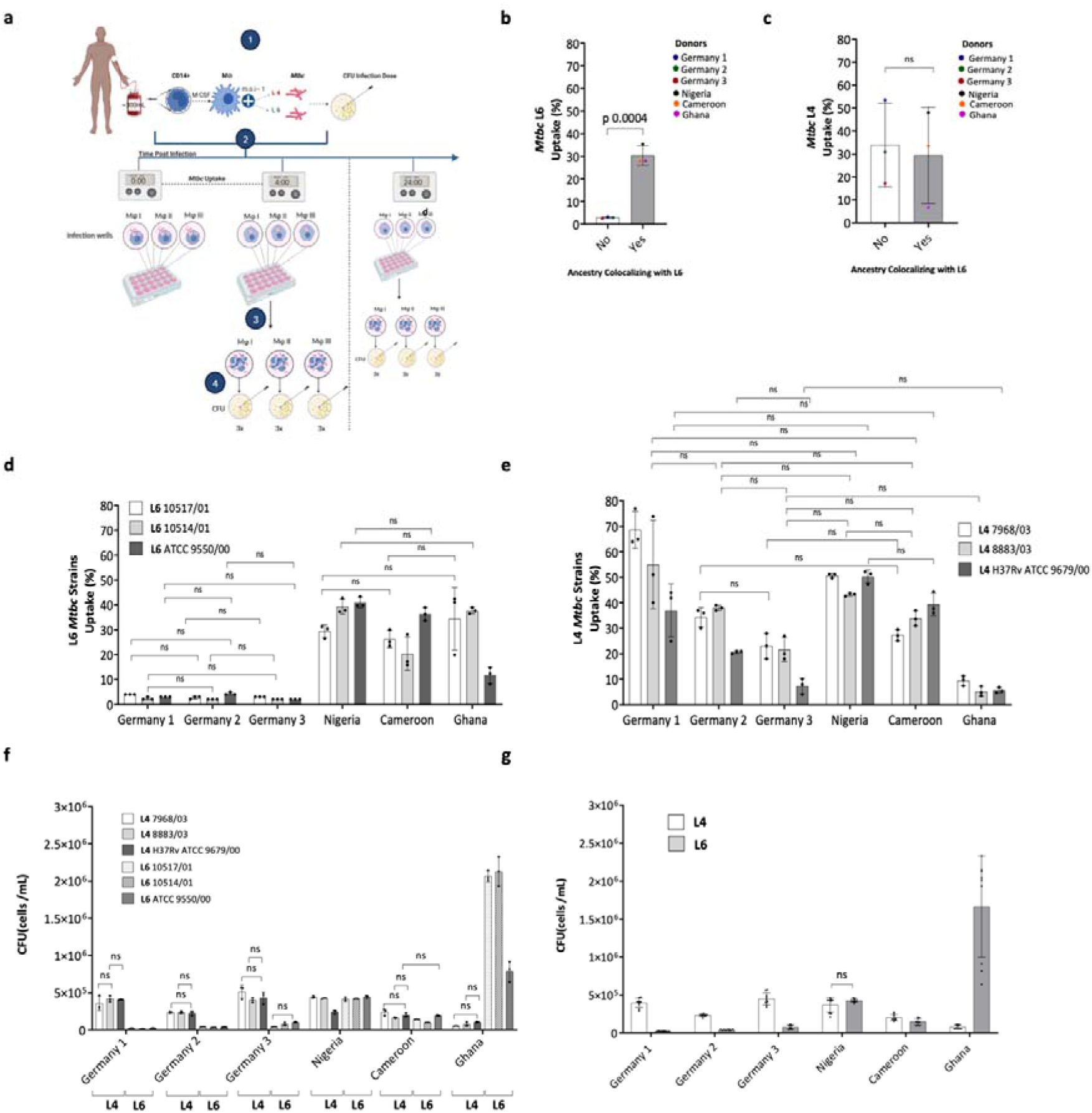
Comparative analysis of phagocytosis and intracellular growth of *M. tuberculosis* complex strains in human macrophages. a. General workflow: 1) Upon obtaining whole blood from healthy donors, CD14+ monocytes were differentiated to macrophages. 2) Macrophages were infected with *M. tuberculosis* complex L4 and L6 strain. 3) Incubation of macrophages with *M. tuberculosis* complex strains for 4h, 24 h, 96h, and 168h. 4) Infected macrophages were lysed, uptake and intracellular growth was assessed by colony forming unit (CFU) assays. (**b-c**) *Mtbc* uptake averaged across three strains of the Euro American Lineage 4 LAM (**b**) and the West African II Lineage 6 **(c)** in blood derived monocyte macrophages (MDMs) from donors of self-reported ancestry that co-localizes with the endemicity of L6. *Mtbc* uptake by MDMs was determined immediately at 4hpi (CFU count at 4hpi × 100%/ CFU count of infection dose) **(d-e)** Phagocytic uptake of two clinical strains and the reference strains of L6 **(d)** and L4 **(e)** lineages in six donors of differing self- reported ancestry. **(f)** *M. tuberculosis* complex intracellular growth of two clinical and the reference strain of L4 and L6 lineages after 24 hours post infection (hpi) in MDMs of donors of different self-reported ancestries (see **Suppl. Figure S6** for 96hpi and 168hpi) **(g)** Intracellular growth at 24 hpi averaged across three L4 and L6 strains by donor. Bars show the mean and standard deviation of the average of CFU values from three infection wells per strain (**Suppl. Figure S3**) and the mean of nine infection wells per lineage respectively. Three CFU measurements from each infection well were performed. MDMs were processed and infected at MOI ~1:1 (0.5 ×10^6^ cells: 0.5 ×10^6^ *Mtbc* bacilli) under the same conditions and in the same laboratory. Statistical results are based on unpaired T-test (b, c, g) or One- way ANOVA with Bonferroni post hoc test correction among donors (d and e), or between strains and H37Rv (f). Mean, standard deviation, and non-significant (ns, p > 0.05) are depicted.

We further investigated whether L4 or L6 strains differed in their intracellular growth kinetics after uptake into macrophages of different ancestries over time (4h, 24h, 96h, and 168h post infection). In macrophages from donors with self-reported ancestry of Nigeria, Cameroon, or Ghana there was no significant difference in growth between L4 and L6 at two of the three time points (**Figure 4f-h, Suppl. Fig S6**). However, growth in macrophages from these donors was higher for L6 compared with L4 strains at 24hpi (comparing average growth across the three strains for each lineage for the Ghanaian donor, p < 0.001, Cameroonian donor p < 0.05, no difference in macrophages of the Nigerian donor) (**Figure 4 e-f**). Conversely, in the macrophages from donors of self-reported German ancestry, L4 strains grew at higher rates than L6 at all the time points tested (p < 0.05 at 24hpi, 96hpi and 168hpi, unpaired T-test and ANOVA with Bonferroni correction) (**Suppl. Figure S6**).

Overall, these results provide evidence that differential transmissibility between allopatric and sympatric pairs may relate to the observed weaker uptake and growth of geographically restricted lineages in allopatric macrophages than increased growth and uptake of these lineages in sympatric macrophages. This suggests that sympatry predominantly manifests in diminished infectivity of geographically restricted *Mtbc* lineages in allopatric hosts.

## DISCUSSION

It is well established that pathogens of genetic lineages of the *Mycobacterium tuberculosis* complex co-localize with human hosts, but it remains unclear if co-localization has any biological effect for infection with *M. tuberculosis*. In this study, we provide the first evidence for a measurable impact of sympatry on the TB epidemic in a controlled study. Our results demonstrate intrinsically reduced transmissibility of strains of geographically restricted *Mtbc* lineages compared with strains of widespread *Mtbc* lineages in three cosmopolitan cities.

While strains of geographically restricted lineages are able to infect sympatric hosts, they display lower transmissibility in allopatric hosts. We validate our epidemiological observations using a macrophage infection model, finding evidence that the reduced transmissibility of geographically restricted strains may relate to lower uptake and growth of *Mtbc* in allopatric macrophages after initial exposure. Our central discovery is that the reduced infectivity of strains of geographically restricted *Mtbc* lineages in allopatric populations is likely responsible for their limited success in driving the global spread of TB compared to strains of modern widespread L2/L4 lineages.

Our results support the theory that geographic co-localization, or sympatry, between pathogen and host can have biological implications and suggests that strains of geographically restricted lineages face a barrier in their ability to migrate to other geographical locations. Patterns of human migration alone are not sufficient in accounting for this geographical spread seen today as evidenced by the little disease burden caused by strains of the geographically restricted *Mtbc* lineages in the Americas. The ecological observations of differing global strain diversity thus do not seem to be only the effect of founder effects and human migration, but a differing human host range of *M. tuberculosis*.

Since bacterial uptake and transmissibility are preserved in sympatric vs allopatric pairs in the analyses presented here suggests a specific role for the host-pathogen pair beyond *Mtbc* lineage alone. Further, TB disease in India, East Africa, and many parts of South East Asia is largely caused by geographically restricted lineages (L1 and L3),^1^ indicating effective transmission of strains of these lineages among local populations. A biological effect of sympatry was also shown for *H. pylori*, a major risk factor of human gastric cancers. A recent study revealed that while colonization with allopatric *H. pylori* strains was correlated with severe gastric cancer, sympatric *H. pylori* infections carry the best prognosis.^28^

Previous attempts at quantifying the biological effect of sympatry in tuberculosis were limited by measuring transmission at the index case level and often did not control for host level variables resulting in inadequate control for social network structure, extent of exposure, and susceptibility to disease. We used a Poisson framework to quantify the number of transmissions among close contacts of each index case. Recent exposure is a substantial risk for all contacts independent of their prior IGRA status (i.e., previous exposure) and TB transmission generally requires significant contact time between a contact and an active case.^29–31^ Our modeling approach and data allowed us to control for contact exposure between lineages and, separately, between sympatric and allopatric contacts because all contacts had a known and similar high-risk exposure to *Mtbc*.

We validated our epidemiological findings in an in vitro macrophage infection assay and found low efficiency in phagocytic uptake in macrophages from donors who did not co- localize with the infecting strain. We cannot test the hypothesis that host ancestry related genes are responsible for phenotypic difference in uptake as we did not obtain human sequence data, yet recent work suggested that up to 9.3% of macrophage expressed genes show ancestry-associated differences in the response to infection.^32^ While more work is warranted to compare more strain-host combinations, including interrogating the host genetics, we had chosen L6 and L4 strains based on their phylogenetic distance and availability of sympatric host macrophage donors. Despite causing a considerable proportion of TB disease in West Africa, L6 strains are associated with reduced virulence in animal models^33^, HIV^34^ and diabetes^35^, and as such could represent a lineage of generally reduced virulence.

Elucidating molecular mechanisms of resistance to *Mtbc* infection will take time, but our findings can be used to improve existing models of TB susceptibility used in public health practice. Although several patient risk factors (e.g., age, sex, HIV, and diabetes) are currently recognized to influence individual risk of TB infection and disease after exposure, they are rarely used quantitatively to support decisions in contact tracing or the initiation of chemoprophylaxis after exposure. Here, we demonstrate that allopatric infections constitute about 52% of total contact exposures to strains of geographically restricted *Mtbc* lineages in low prevalence settings and influence the odds of infection to a similar extent as recognized risk factors such as index case smear grade. The incorporation of *Mtbc* sympatry for geographically restricted *Mtbc* lineages as well as host factors into the baseline assessment of risk using statistical models may help refine the use of chemoprevention and guide contact tracing activities. Moreover, identifying the components of Mtbc genetic variation responsible for host specificity can inform vaccine design to elucidate pathogen targets of host susceptibility.

The investigation of the effects of sympatry between humans and *Mtbc* strains requires a diverse cohort that ensures both infections with sympatric and allopatric strains are adequately represented in the sample. This study was enabled by the availability of robust TB case data, pathogen WGS, and contact tracing data and illustrates that these data can successfully be repurposed and pooled across several public health systems. Yet, this study was not without limitations. The TB index case and contact information relied on social contact tracing that may be subject to recall bias. Information on contacts of TB cases is not routinely collected in Hamburg and Amsterdam and as a result some key risk factors of TB susceptibility (contact HIV, contact diabetes) were not available. The analysis of secondary active TB relied on modeling rather than epidemiological linkage which may include false positive transmission links, and some contacts of TB index cases may not be visible to the public health departments due to contact relocation between cities or states. We used country of birth, or in case of minors, the parent’s countries of birth, as a surrogate for ancestry. As self-reported ancestry is a poor proxy for human genetics our approach did not allow us to directly measure host-genetic adaptation in this study. We performed several sensitivity analyses to address these limitations. We randomly assigned the sympatry label for 14% of our sample (the proportion of Dutch residents who are foreign-born, Statistics Netherlands) to account for ancestry misclassification. Estimated transmission links were only accepted based on sample size and support value thresholds that were shown to provide robust effect estimates in our study and other work.^36^ We built models of transmission at contact level in a subset of data where HIV and diabetes information was available, as these are important effect modifiers of TB susceptibility. Finally, the differing uptake efficiencies by macrophage donor – pathogen strain combinations may impact the growth kinetics of *Mtbc* strains. To enable comparability, all macrophages were infected with the same amount of *Mtbc* bacilli, and the infection dose was quantified by plating the inoculum on agar plates for CFU counting.

In summary, we demonstrate an epidemiological impact of sympatry that manifests in lower infectivity of geographically restricted *Mtbc* lineages in allopatric host-pathogen combinations. This suggests that strains of widespread lineages are likely to increase in prevalence relative to strains of geographically restricted lineages in a progressively globalized world. Future work should aim to understand the molecular underpinnings of allopatric versus sympatric infections.

## Data Availability

All data produced in the present study are available upon reasonable request to the authors

## Data availability

Bioprojects and accessions for each fastq file have been requested and will be available in Supplementary table S8.

## Acknowledgements

We thank Pascal Lapierre, Han de Neeling, and Thomas Kohl for transferring sequence data, Henrieke Schimmel and Erika Slump for help with data linkage of the Dutch dataset, the Dutch TB Registry (NTR commissie) as well as all study participants. We thank the Wadsworth Center Applied Genomics Technology Cluster for whole genome sequencing.

## Author contributions

MRF, and MIG conceived the idea for the epidemiological analysis. SN, SH, FJPL conceived the idea for the *in vitro* experiments. MRF supervised the project. MIG performed data curation and data analysis. MIG and MRF wrote the first draft. FJPL performed data curation and data analysis. RVJr and DK carried out data analysis. LT, PK, RD carried out data acquisition. VE, KM, JSM, SH, DvS, SA, SN supervised data acquisition and curation. WS and BM critically reviewed the drafts. All authors reviewed the draft and assisted in the manuscript preparation.

## METHODS

### Study design, participants, and associated metadata

We harmonized TB surveillance data from public health departments tracing TB in three large cities Amsterdam, Hamburg, and New York City, where TB is reportable to the respective public health agencies. The data consists of clinical characteristics and pathogen whole genome sequence data of TB index cases, as well as information on their close contacts including exposure time (close contact versus non-close contact), demographics, relevant comorbidities, and Interferon Gamma Release Assay (IGRA) results. We detail each TB surveillance sample by site below:

Amsterdam and the Netherlands: The National Institute for Public Health and the Environment (RIVM) in Bilthoven, The Netherlands, contains the Mycobacterial reference laboratory for the diagnosis of all TB cases in the Netherlands. Since retrospective data in this study was de-identified by name, WGS data from the RIVM and clinical and demographic data for patients were linked based on sex, date of birth, year of diagnosis and postal code. All notified *M. tuberculosis* culture-positive cases from the city of Amsterdam between December 2015 and December 2019 and their contacts were included. In addition, we obtained information on all active cases diagnosed elsewhere in The Netherlands in this period. In the case of patients with multiple isolates, only the isolate with the earliest date of diagnosis was included. Close contact was defined as daily contact in a room of in total ≥48 hours or ≥6 hours in a car per week.

Hamburg: We obtained data from the Public Health Office Hamburg, Germany for TB index cases from 2008 through 2017. Sequencing of Hamburg cases was performed at the National Reference Center for Mycobacteria at the Research Center Borstel. Close contact was defined as a contact time of in total ≥8 hours.

New York City: The New York City Department of Health and Mental Hygiene notifies, and traces contacts of all infectious TB index cases. We obtained clinical and demographic data of TB index cases diagnosed between 2016 and 2020 and their contacts. Pathogen WGS is performed at the New York State Wadsworth Center in Albany, NY, for culture-positive cases. In the case of patients with multiple isolates, only the isolate with the earliest date of diagnosis was included. Close contact was defined as a contact time of ≥8 hours per week.

We compared characteristics of index cases, and their contact group networks to explore comparability of contact tracing activity across the three cities’ public health departments. Across the three cities, index case age positively correlated with mean contact group age (pooled Pearson’s correlation coefficient 0.4 [CI_95_ 0.39 - 42], and the count of TB infected close contacts correlated moderately with the contact group size (Pearson’s 0.5 [CI_95_ 0.53 – 0.56]) (**Figure S7a-b**). We excluded extrapulmonary TB cases from the source case analysis due to their recognized significantly lower infectiousness (n = 903/5,256,17%). We excluded strains not typed as *Mtb* sensu stricto or *M. africanum* (i.e., *M. canettii* or *M. bovis*, n = 132/5,219, 0.03%). Among contacts, we excluded those without blood test result for *Mtbc* infection (Interferon Gamma Release Assay, or IGRA, 12%, 3,483/28,889), excluded contacts not considered close (42%, 12,228/28,889) and contacts not linkable to an index case (17%, 4,891/28,889). Our rationale for excluding contacts without significant exposure time (<8 hours) independent of exposure location (household versus work or leisure) is based on previous evidence that households account for <20% of TB transmission^1^, and logistic regression models indicating a significant effect of >8h exposure on IGRA positivity (Odds Ratio 2.24, 95% Confidence Interval 2.06 to 2.82, Wald p-value <0.001) (Suppl. Table S8).

### Index case and contact data linkage

Demographics, comorbidity information, and data on social risk factors for TB were extracted from the three public health databases. The investigated contacts were considered to have TB infection if their IGRA values were above 0.35 IU/ml based on existing test specifications. Contacts were linked to TB index cases by epidemiological investigation. This linkage of contacts and active cases is coded as a unique identifier that was subsequently used to compute aggregated contact group information (count and percent of high-incidence country birthplaces) per index case. Conversely, index case information was linked to the contact data (age, sex, sputum microscopy, co-morbidities) as these were important covariates to control for in the modeling.

### Ethical approval

The study is approved by the Institutional Review Boards at Harvard Medical School (IRB19- 1910), the University of Lübeck (AZ-19-071), the New York City Department of Hygiene and Mental Health (20-078), and the Registration Committee of the Netherlands Tuberculosis Register NTR (11-2019).

### Phenotypic drug resistance

At RIVM (The Netherlands), the Research Center Borstel (Germany) and the New York State Wadsworth Center phenotypic drug susceptibility testing was performed with the MGIT 960 method using the World Health Organization recommended critical concentrations of 1mg/L for rifampicin and 0.1 mg/L for isoniazid.

### Whole Genome Sequencing and analysis

At the time of writing, whole genome sequencing of all culture positive TB cases is routinely performed in the Netherlands and New York State since 2016.

Isolates from the Netherlands were sequenced at the RIVM Bilthoven as follows: genomic DNA was extracted from positive MGIT culture tubes (Becton Dickinson, NJ, USA) using the QIAamp DNA mini kit (QIAGEN GmbH, Hilden, Germany) and sequencing was performed on Illumina HiSeq2500 with 2 × 125bp reads following Nextera XT library preparation.

At the Wadsworth Center of the New York State Department of Health, genomic DNA was extracted from MGIT culture tubes (Becton Dickinson, NJ, USA) using the InstaGene/FastPrep method as previously described.^2^ Paired-end 250-bp DNA sequencing was carried out using the Illumina MiSeq platform (Illumina, San Diego, CA, USA) after Nextera XT library preparation.

Patient isolates from Hamburg were sequenced for a previous research project at the Research Center Borstel as follows:^3^ Genomic DNA was extracted from solid Löwenstein Jensen culture tubes using the Cetyltrimethylammonium Bromide (CTAB) method. Sequencing libraries were constructed from genomic DNA using a modified Illumina Nextera protocol for 2 × 151 bp reads on the Illumina NextSeq 500 (Illumina, San Diego, CA, USA).

Raw sequencing reads across the three sites were pooled and analyzed on Harvard Medical Schools High Performance Cluster O2. Analysis and quality control was performed using the same bioinformatics pipeline. Reads were first trimmed to remove low quality bases and adapter sequences using *fastp* v0.20.1.^4^ The trimmed reads were aligned to the reference genome H37Rv (accession NC_000962.3) using *BWA mem* v0.7.17 (parameters: default) ^5^. Duplicate reads were removed with *Picard* v2.25.1 (http://broadinstitute.github.io/picard/). We called single nucleotide substitutions (SNSs) and indels with *Pilon* v1.24 (parameters: default) requiring confidence base calls (*pass* filter).^6^

### Variable position alignment and phylogenetic analysis

To analyze the genetic variation in our sample we created a genotypes matrix based on the variant call format files. We detected SNS sites at 347,471 H37Rv reference positions (of which 9,641 SNSs were not biallelic) among our global sample of 5,219 strains. We constructed a 347,471×5,219 genotypes matrix (coded as 0:A, 1:C, 2:G, 3:T, 9:Missing) and filled in the matrix for the allele supported at each SNS site (row) for each isolate. We excluded 7,406 SNS sites that had an Empirical Base-level Recall (EBR) score of <0.9,^7^3,555 SNS sites in mobile genetic elements (e.g., transposases, integrases, phages, or insertion sequences), and 933 SNS sites located in overlapping coding sequences. We next dropped 10,169 SNS sites that are missing in >10% of our strains. An additional 133 SNS sites were dropped because the minor allele is present in <1 isolate. If a base call at a specific reference position for an isolate did not meet the filter criteria that allele was coded as missing. Our filtered genotypes matrix had dimensions of 325,275×5,219 SNSs representing 325,275 SNS sites across 5,219 strains. The phylogenetic lineage was determined using a refined lineage-calling scheme based on 96-SNSs with *fast-lineage-caller*.^8^

To infer the population structure of our strains we merged the variant call format files using *bcftools* v1.12^9^ and removed regions with repetitive sequence content with low EBR scores,^7^ mobile genetic elements, coding for genes involved in antibiotic resistance as well as all invariant sites. A multiple sequence alignment of 297,061 characters found in 5,087 *Mtbc* L1- L6 strains was created using *vcf2phylip* (v1.5, https://doi.org/10.5281/zenodo.1257057) and fed to *iqtree* v2.1.2 for phylogenetic inference.^10^ The final maximum likelihood tree was inferred using a general time reversible model of nucleotide substitution, a Gamma model of rate heterogeneity, empirical base frequencies (GTR+F+R2) and 1000 ultrafast bootstrap replicates. The tree was annotated using iTol.^11^

### Definition of clusters based on genetic distance

*Mtbc* strains from patients linked by transmission using epidemiological data rarely differ by more than 5 SNSs.^3,12^ To define clustered strains in our dataset, we first excluded the small number of non-biallelic SNSs to simplify distance calculation. Of the 325,275 SNS sites, 316,274 were biallelic, i.e., had only a single alternate allele among our strains. We calculated the pairwise distance as the Hamming distance of SNSs across the total 5,219 strains and subset this matrix for to obtain one pairwise distance matrix for each *Mtbc* lineage and city.

We next performed graph-based clustering using the *gengraph* function implemented in the adegenet R package v2.1.5^13^ for each *Mtbc* lineage and city specific pairwise distance matrix as we were only interested in transmission within each city. We used a 12 SNS cut-off as possible transmission with single linkage clustering, i.e., strains in a cluster differed at most 12 SNSs from at least one other isolate in the cluster.

### Index case inference and infector/infectee relationships

We used the R package phybreak v0.5.2 to identify the infector case for each active TB case in a cluster.^14^ Cases that were not in clusters were assumed to not have secondary active TB cases. (If contacts of TB cases moved to a different city and developed secondary active TB this would not have been detected using this approach.) Phybreak identifies infector- infectee relationships by combining models for transmission, case observation, within-host pathogen dynamics, and mutation assuming that all cases of a transmission cluster have been sequenced. The most likely transmission tree is sampled from the posterior distribution through Bayesian inference and Monte Carlo Markov Chain sampling. The priors for the mean and shape of the sampling interval (time between infection and sampling of a case) and generation interval (i.e., time between infection of a primary case and a secondary case by this primary case) were assumed to follow a gamma distribution as in previous work ^15,16^. We ran five independent MCM chains with 10,000 cycles of burn-in and 50,000 sampling cycles for each cluster. We dropped each estimated infector-infectee relationship where the effective sample size was <200 which can occur if sampling dates occur within a short period of time. We excluded active cases that did not have an infector with a posterior probability (PP) of at least 0.5 from further analysis after a sensitivity analysis evaluating different PP values in the transmission models (**Suppl. Fig S8**).

### Statistical analysis and models of transmission

The python libraries *pandas* v1.3.5^17^ and *numpy* v1.21.5^18^ were used to manipulate epidemiological data. All statistical analyses and visualizations were performed in R v4.1.2^19^ using libraries included in the *tidyverse* v.1.3.1.^20^ Effect estimates of regression analyses were visualized using the *sJPlot* package 2.8.10. Missing values for the covariates HIV coinfection and alcohol were imputed using a bootstrapped logistic regression model implemented in the package *mice* v3.14.0^21^. We coded age into brackets (0 to 14, 15 to 24, 25 to 34, 35 to 44, 45 to 54, 55 to 64, 65 to 74, and 75 and above).

We used a multivariate Quasi-Poisson rate regression model (Eq1) to identify predictors of the number of secondary active TB cases or *Mtbc* infections among close contacts. We ensured there was no collinearity between predictor variables (Suppl. Figure S8). The group size of close contacts per index case was used as the offset to denote the exposed group in the Poisson regression. Adjusted incidence rate ratios are presented with 95% confidence intervals.

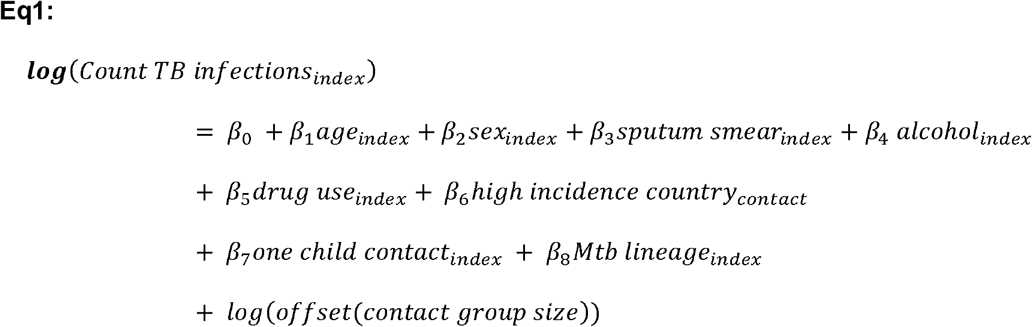

Generalized estimating equations (GEE) were used to validate the Quasi-Poisson models and to quantify the effect of host-pathogen sympatry on TB infection for each close contact individually (Eq2). The *gee* package v4.13-20 was used for these analyses with the option ‘exchangeable’ to imply that any two observations per contact group are equally correlated without correlation between observations from different contact groups ^22^. This allowed to adjust the effect estimates for correlations in the data. Adjusted odds ratios and 95% CI are presented.

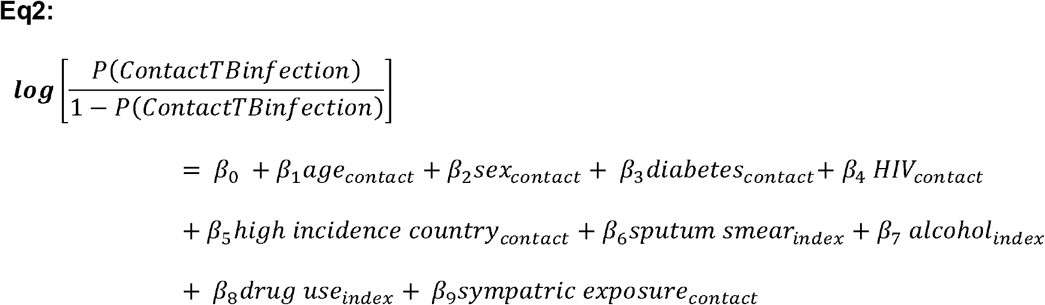

Frequencies or proportions of binary variables were compared using Chi Squared, binary to continuous variables were compared using the Wilcoxon Rank Sum test, and continuous data was compared using ANOVA or t-tests. All statistical tests were two-sided and a *P* value cut-off of 0.05 corrected for multiple testing with the false discovery rate.

### Host infectiousness scores

The adjusted estimates from the multivariate Quasi-Poisson regression model were each multiplied as weights to calculate each index case’s infectiousness score, similar to previous work on the propensity to propagate.^23^

### Human macrophage donors

Blood donation was approved by the ethics committee of the Research Center Borstel (RCB) (Borstel, Germany) and the University of Luebeck (Luebeck, Germany) (AZ-19-071). A total of ~ 300 ml of whole blood (heparin- anti-coagulated) was obtained via venepuncture by the blood service unit at the RCB. Human healthy donors self-reported ancestries from Germany, Nigeria, Ghana, and Cameroon with a medical check-up no longer than two years ago.

### *Mtbc* strains and culture conditions

A panel of six *Mtbc* strains representing the modern and ancient human-adapted clades 1 and 2 (L4 and L6) was selected from a reference collection at the RBC. Strains were recovered from clinical samples or provided by the American Type Culture Collection (ATCC). Strains were characterized by drug susceptibility testing and sequencing as previously described.^24,25^ *Mtbc* strains were grown on Loewenstein-Jensen (LJ) medium and further passaged in 10 ml of mycobacteria liquid media (MLM): Middlebrook 7H9 broth (Becton Dickinson) supplemented with 10% oleic acid albumin dextrose catalase (OADC), 0.2% glycerol, 0.05% Tween 80. *Mtbc* bacilli were grown until mid-log phase at an Optical Density (OD)_600nm_ of 0.5 at 37 °C, harvested, and stored as stocks in MLM at -80 °C. Aliquots were finally titrated by Colony-Forming Units (CFU) using Middlebrook 7H10 agar (Becton Dickinson) plates supplemented with 10% heat-inactivated bovine serum and cycloheximide (10 mg/ml).

### *In vitro* generation of macrophages

Blood-derived human monocytes were obtained by density gradient centrifugation and separation with anti-human CD14 antibodies (Milteny Biotec, Germany) following the manufacturers instructions. Isolated monocytes had a purity and viability greater than 85- 90% which was assessed by flow cytometry and electronic current exclusion technology with CASY. Monocytes were seeded in 24 well plates at 0.5×10^6^ cells in 500 µl of Macrophage Culture Media (MCM): RPMI 1640 (Biochrom), penicillin/streptomycin 1% (Biochrom), L-glutamine 1% (Biochrom) with AB + heat-inactivated human serum 1% and further differentiated into monocyte-derived macrophages (MDM) after seven days of culture in the presence of 10 ng/ml of human recombinant macrophage colony stimulating factor (M-CSF). Cells were incubated at 37°C /5% CO_2_, and MCM was changed every three days until cells achieved ~ 80% confluence.

### Cell infection assays

CFU-titrated *Mtbc* stocks were used to prepare the bacterial suspension for infection experiments in macrophage infection media consisting of 1% L-Glutamine and 10% FSC in RPMI medium (all Biochrom). For growth experiments, each experimental setup comprised four plates for different time points (4 h, 24 h, 96 h, and 168 h post-infection (hpi)). **0.5 × 10**^**6**^ *Mtbc* bacilli per 500 µl Macrophage infection media were used to infect **0.5 × 10**^**6**^ MDM per well. Macrophage infection media without bacteria was used as infection control. Upon infection, plates were incubated at 37 °C in 5% CO_2_ for 4h. We quantified CFUs of the inoculate to be able to compare the infection dose. The percentage of *Mtbc* uptake was calculated as colonies count at 4hpi*100%/colonies count of infectious dose. Each time point included three replicates per *Mtbc* strain and three non-infected controls. In all *in vitro* cell infection experiments, the number of intracellular *Mtbc* bacilli was determined by CFU assays four hpi and at these different time points: 24 hpi, 96 hpi, and 168 hpi. Cells were first lysed with sodium dodecyl sulfate (SDS) 0.05% and lysate was then plated in triplicates onto Middlebrook 7H10 agar plates for CFU counting.

Three independent infection experiments of donors with self-reported ancestry of Germany were performed representing a total of nine technical replicates of infection of which each was plated three times for CFU quantification. Due to insufficient cell yield two experiments compromised two donors each. Of these experiments, in one donor (Germany 2), L4 and L6 strains were assayed in macrophages of one single donor while the other two experiments (Germany 1 and 3) comprised two donors. Of these, the first donor was infected with L4 and H37Rv, the second donor with L6 and H37Rv. These data are represented together in one figure as we detected no significant differences in H37Rv uptake, growth, and inter-intra host variation among donors (**Suppl. Figure S9 a-h and Figure S10 a-f)**. Three independent infection experiments were conducted including donors from Nigeria, Cameroon, and Ghana. Each compromised one single donor infected with L4 and L6 (including nine technical replicates of infection, and 27 CFU technical replicates per strain).

Supplementary Information is available for this paper.

Correspondence and requests for materials should be addressed to Matthias I Gröschel or Maha R Farhat.

