## Supplemental Figures and Tables for "Host-pathogen sympatry and differential transmissibility of *Mycobacterium tuberculosis complex*": 2023-02-15_supplement.pdf

Supplementary figures and tables

### SUPPLEMENTARY MATERIAL

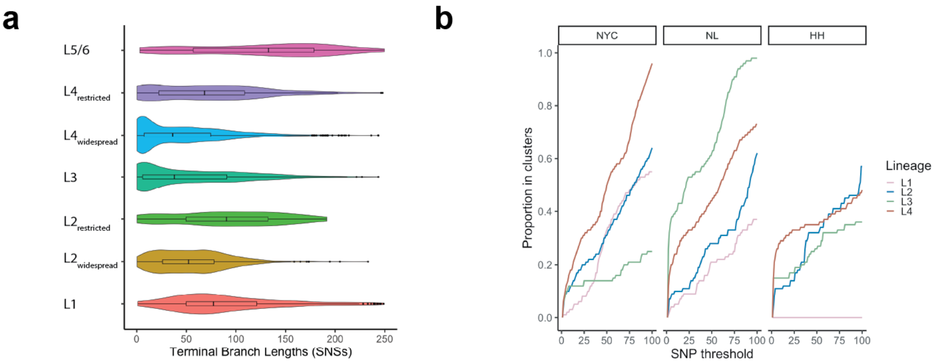


**Legend for Suppl. Figure S1: Genetic characteristics of *M. tuberculosis* complex strain.** a) Violin plot of the terminal branch lengths of the included *Mtbc* genetic lineages. Medians and Interquartile Ranges are plotted on the figure for each Lineage b) Proportions of strains in clusters based on several different Single Nucleotide Substitution thresholds by genetic lineage and site. L = Lineage, SNP = Single Nucleotide Polymorphism, NYC = New York City, NL = The Netherlands, HH = Hamburg.

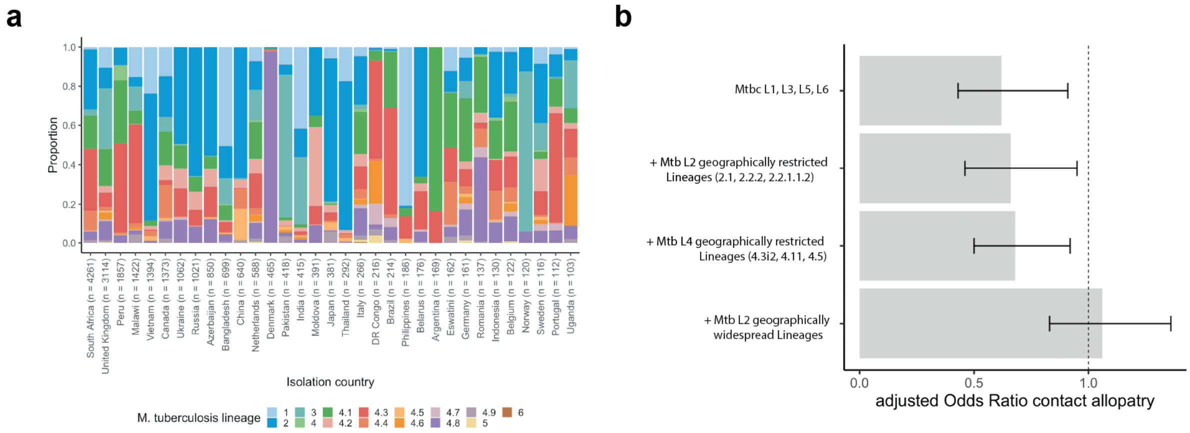


**Legend for Suppl. Figure S2: Relationship of index case self-reported ancestry and *M. tuberculosis* complex lineage.** a) Bar plot detailing the proportions of isolation country and *Mtbc* lineage in a global sample of 25,243 strains. b) Adjusted odds ratios estimated for the variable contact allopatry using different co-localization or sympatry assumptions from multivariate Generalized Estimation Equation models (see Figure 3f in main text).

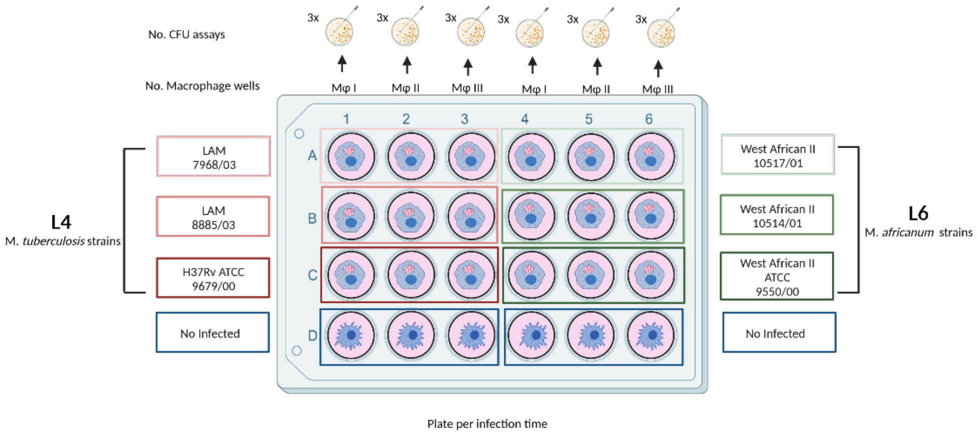


**Legend for Suppl. Figure S3: Experimental Design of in vitro uptake and growth experiments of different pathogen donor combinations**. Experimental setup of a representative plate: One independent experiment comprised four plates of 24 wells for the four infection time points evaluated per donor, each plate comprised control wells with non-infected macrophages, and wells with macrophages infected with the six or four strains of a given lineage (including the reference strain H37Rv ATCC 9679/00). Each macrophage-starin combination was performed in triplicates well replicates, and three colony forming unit (CFU) measurements were done per well for a total of 9 CFU replicates per strain and time point assessed. The figure was designed with Bio-Render.

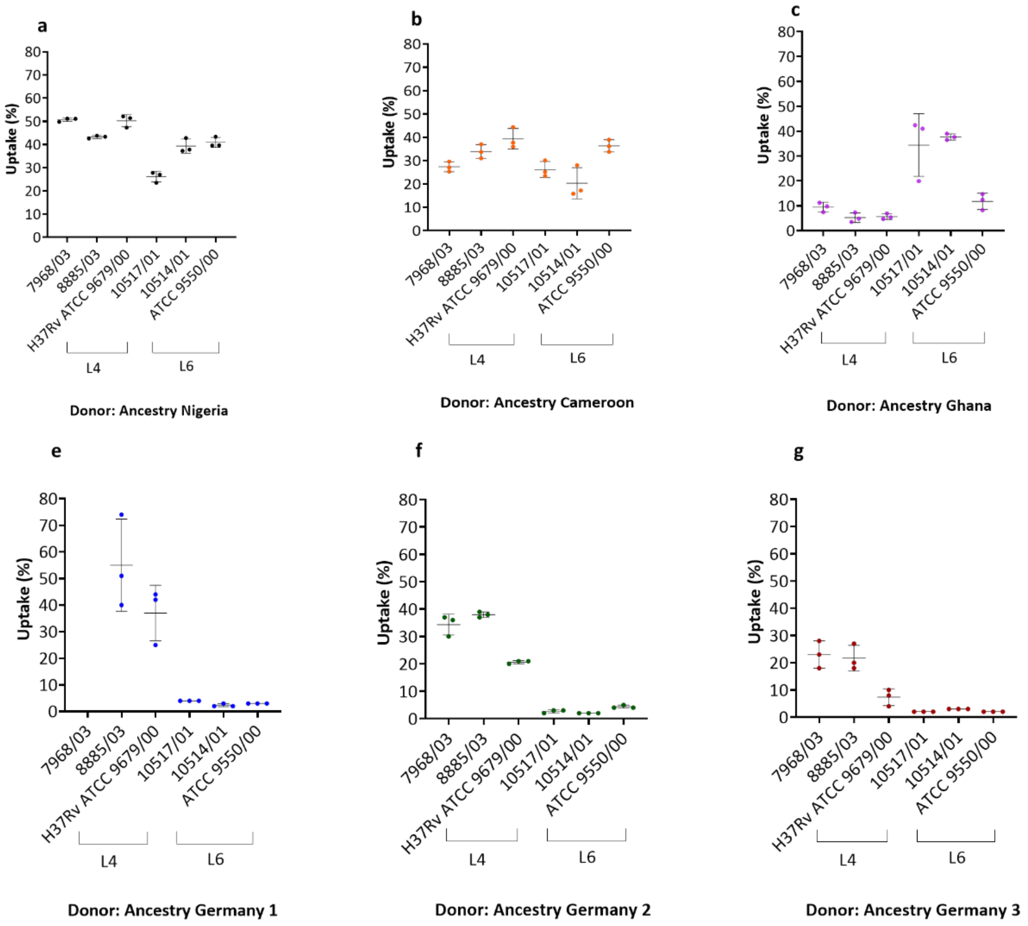


**Legend for Suppl. Figure S4: *Mtbc* Uptake Intra Host Variation Analysis.** Human donor self-reoprted ancestries: Nigeria (**a**), Cameroon (**b**), Ghana (**c**), Germany 1 (**d**), Germany 2 (**e**), and Germany 3 (**f**). Macrophages were infected at MOI ~1:1 (0.5 x$10^6$ cells: 0.5 x$10^6$ *Mtbc* bacilli) and the numbers of intracellular bacteria (CFU) were determined immediately after uptake at 4h post-infection. Uptake percentage (y-axis) and *Mtbc* strain (x-axis). A single dot represents the mean of three CFU measurements per infection well therefore three dots represent three infection well replicates per strain assayed (n=3). Mean and standard deviation are depicted in the figures. Abbreviations: *Mtbc*, *Mycobacterium tuberculosis* complex; L4, Lineage 4; L6, Lineage 6; CFU, Colony Forming Unit; MOI, Multiplicity of Infection.

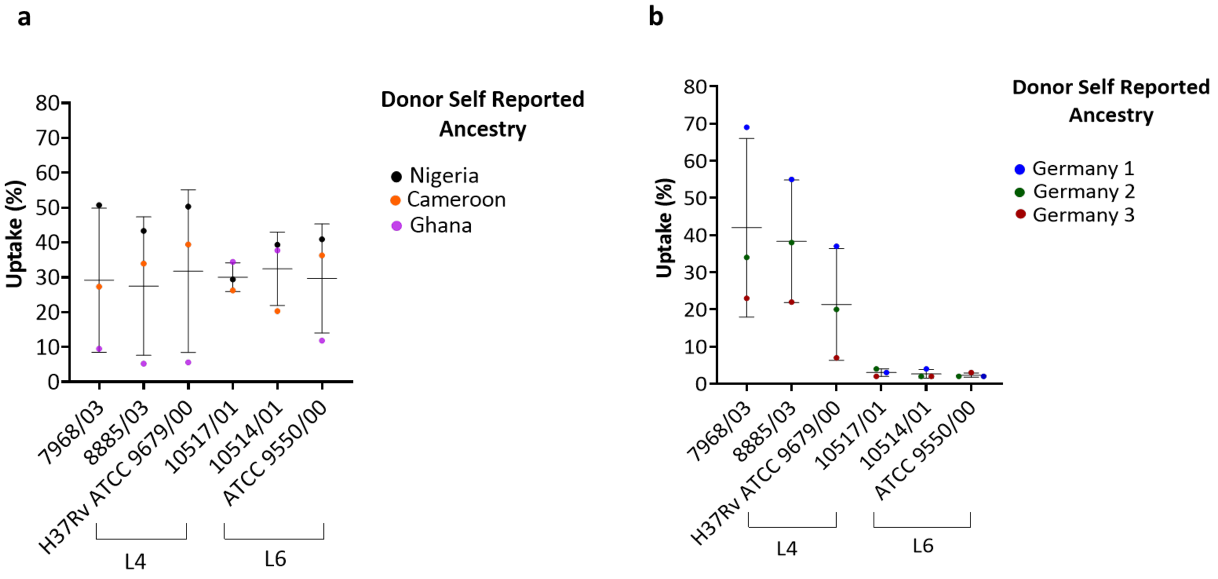


**Legend for Suppl. Figure S5: *Mtbc* Uptake Inter Host Variation Analysis. (a)** Donor Self-Reported Ancestry: Nigeria, Cameroon and, Ghana, and (b) Germany 1, Germany 2, and Germany 3. Macrophages were infected at MOI ~1:1 (0.5 x$10^6$ cells: 0.5 x$10^6$ *Mtbc* bacilli) and the numbers of intracellular bacteria (CFU) were determined immediately after uptake at 4hpi. Uptake percentage (y-axis) and *Mtbc* strain (x-axis). A single dot represents the mean of three infection macrophage well (n=3) per strain per donor. Mean and standard deviation are depicted in the figures. Abbreviations: *Mtbc*, *Mycobacterium tuberculosis* complex; L4, Lineage 4; L6, Lineage 6; CFU, Colony Forming Unit; MOI, Multiplicity of Infection; h, hours.

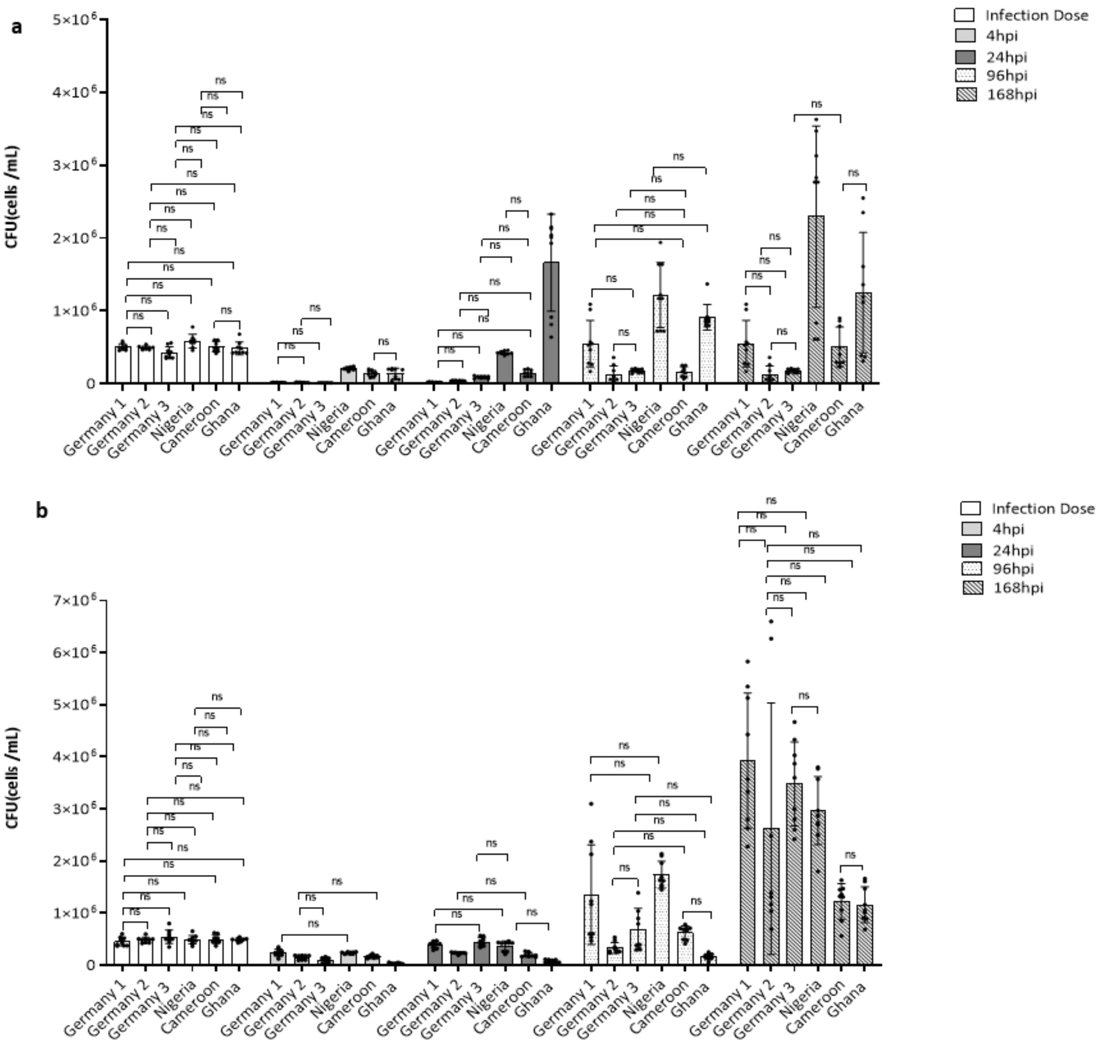


**Legend for Suppl. Figure S6: Comparative analysis of intracellular growth of *M. tuberculosis* complex strains in human macrophages.** Intracellular growth of *Mtbc* L6 (**a**) and L4 (**b**) at 4hpi, 24hpi, 96hpi, and 168 hpi in monocyte derived macrophages (MDMs). *Mtbc* growth was assessed by colony forming units (CFU). Each lineage represents the average of three representative strains. The mean and standard deviation of CFU values from three infection wells (n=3) per strain and nine infection wells (n=9) per lineage and donor respectively (See **Suppl. Figure S3**) are shown. MDMs from donors of each self-reported ancestry were infected nine times with the respective strains of the two lineages and intracellular growth was measured twenty-seven times using CFU. The mean of CFU values per infection well is presented as a single dot on the figures. Statistical results shown in the figures are based on One- way ANOVA multiple comparisons with the Bonferroni post hoc test among donors within a given time point. Mean, standard deviation, and only no statistical differences are depicted in the figures (ns >0.05). Abbreviations: *Mtbc*, *Mycobacterium tuberculosis* complex; L4, Lineage 4; L6, Lineage 6; CFU, Colony Forming Unit; MOI, Multiplicity of Infection; h, hours.

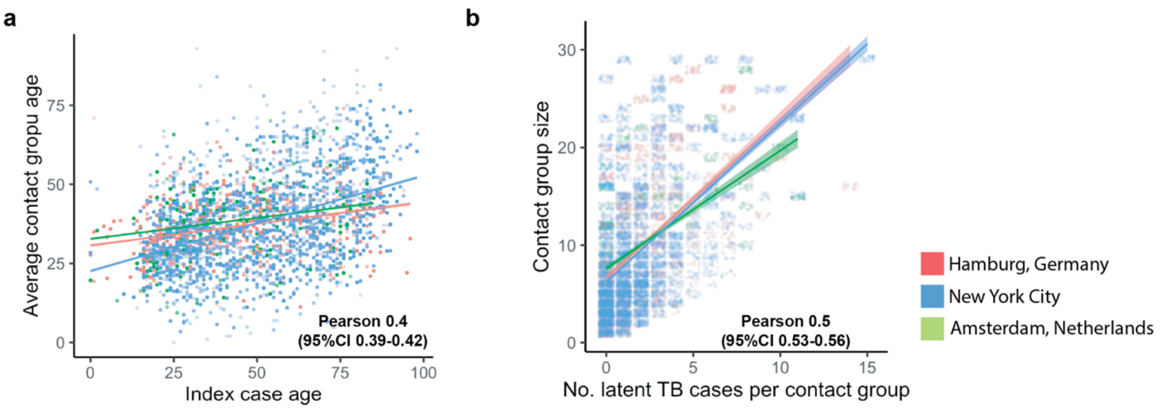


**Legend for Suppl. Figure S7: Comparison of index case and contact group characteristics**. a) Dot plot of the index case age (x-axis) versus mean contact group age (y-axis) for each of the included cities; b) Dot plot of the No. of *M. tuberculosis* infections per contact group (x-axis) and the size of the contact group (y-axis).

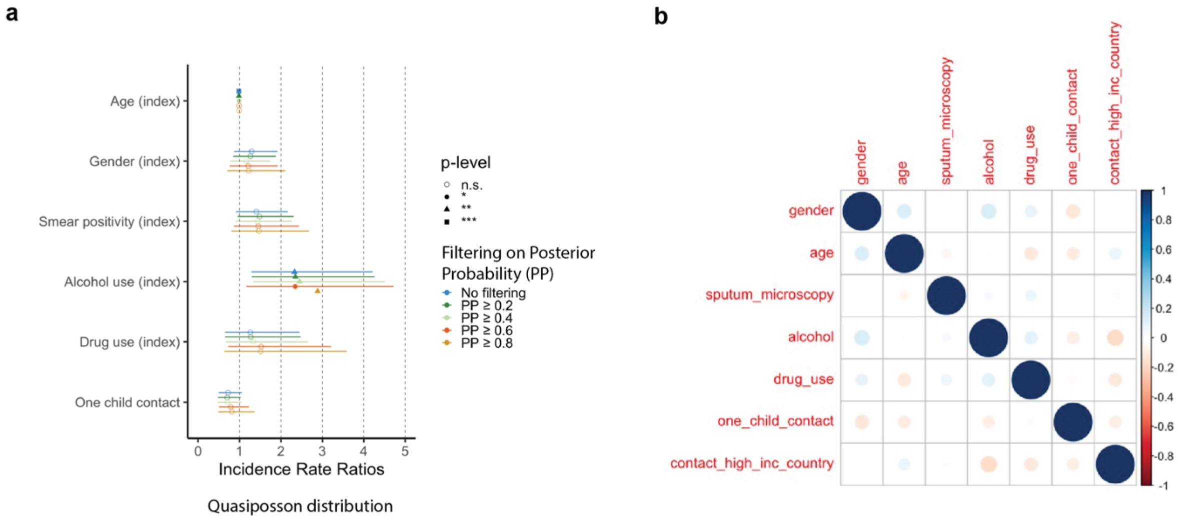


**Legend for Suppl. Figure S8: Impact of different support value thresholds on effect estimates of transmission. a)** Forest plot of effect estimates from a quasi-Poisson count regression model predicting the count of secondary active tuberculosis cases for each index among his close contacts. Several different posterior probability values were evaluated after inference of transmission relationships using Phybreak (1). **b)** Analysis of collinearity between predictor variables using Pearsons correlation coefficient.

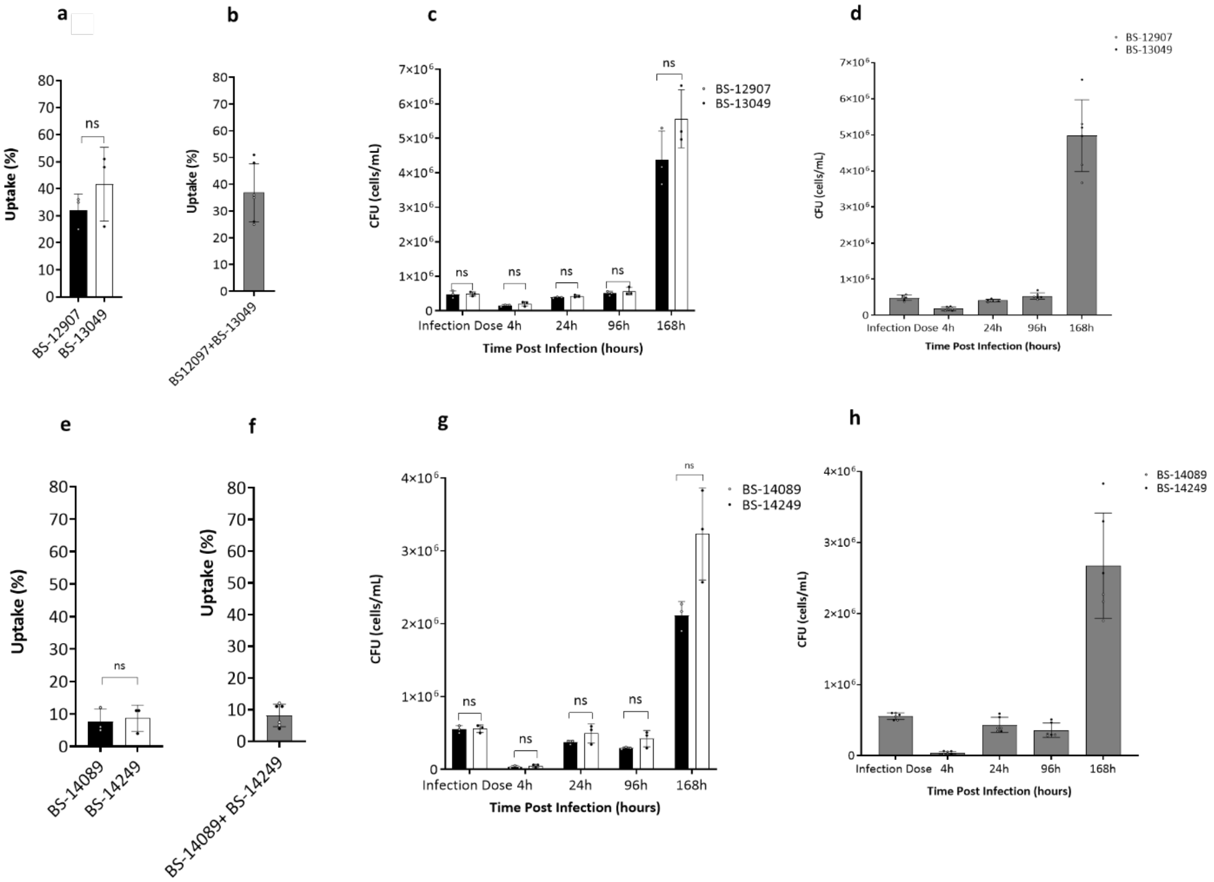


**Legend for Suppl. Figure S9: Comparison of uptake and intracellular growth of *M. tuberculosis* H37Rv ATCC 9679/00 laboratory reference strain among two donors of German self-reported ancestry comprising donors Germany 1 and Germany 3.** Top panel represent experiments BS-12907 and BS-13049 for Germany 1 (**a-d**): uptake per donor (**a**), uptake average among donors (**b**), intracellular growth upon infection per donor (**c**) and, intracellular growth average among donors upon infection (**d**). Bottom panel represent experiments BS-14089 and BS-14249 for Germany 3 (**e-h):** uptake per donor (**e**), uptake average among donors (**f**), intracellular growth upon infection per donor (**g**) and, intracellular growth average among donors upon infection (**h**). Blood monocyte derived macrophages were infected at MOI~1:1 ($0.5x10^6$ cells: $0.5x10^6$ bacteria) and the numbers of intracellular bacteria (CFU) were determined immediately after uptake at 4hpi, 24hpi, 96hpi and 168hpi post-infection. Three infection wells per strain (n=3) and per time point were assayed per donor and three CFU measurements from each infection well were performed. Statistic results were determined by T-test among experiments. No statistical differences are depicted in the figure (ns >0.05). Error bars represent the standard error of the mean of three CFU values from three infection wells per strain and time point. Three CFU measurements from each infection well and infection dose were performed. Abbreviations: CFU, Colony Forming Unit; MOI, Multiplicity of Infection; hpi, hours post infection.

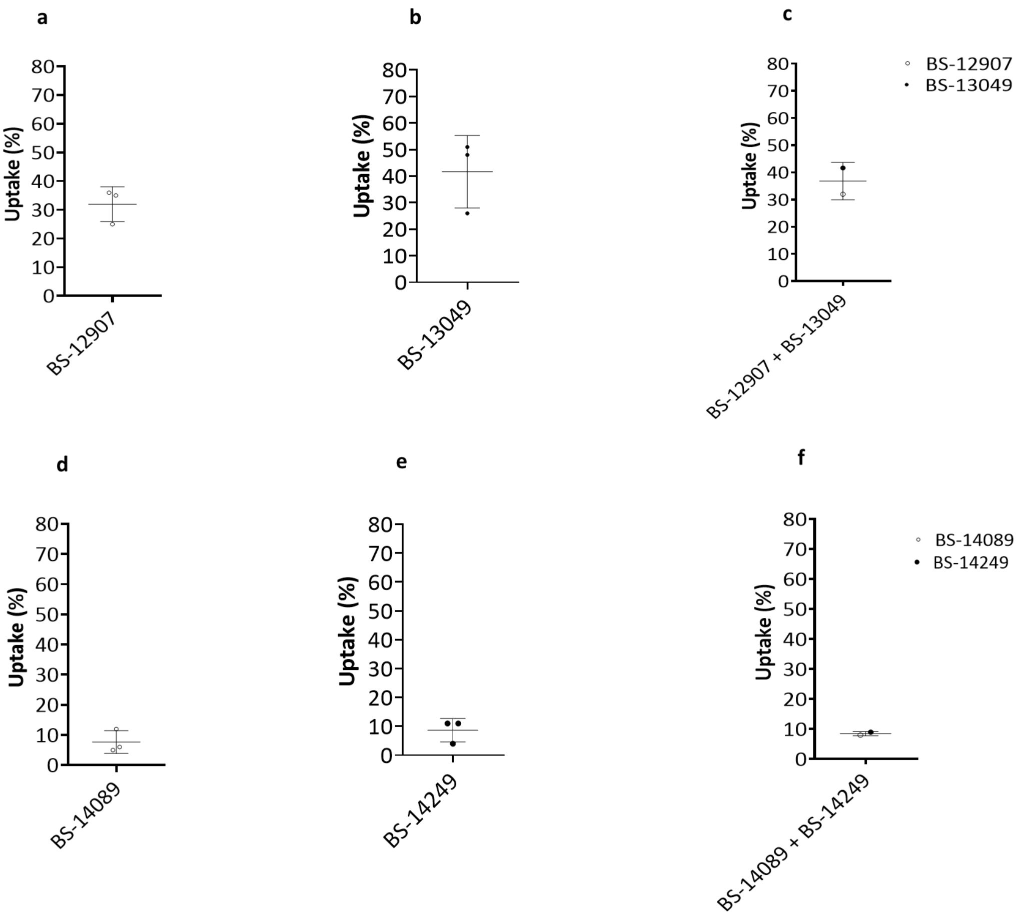


**Legend for Suppl. Figure S10: Host Variation Analysis of *M. tuberculosis* H37Rv ATCC 9679/00 uptake among two experiments comprising Germany 1 and Germany 3 experiments.** Top panel represent donor BS-12907 and BS-13049 for Germany 1 (**a-c**): Intra host variation (**a-b**) and inter host variation (**c**). Bottom panel represent experiments BS-14089 and BS-14249 donors for Germany 3 (**d-f**)**:** Intra host variation (**d-e**) and inter host variation (**f**). Macrophages were infected at MOI ~1:1 (0.5 x$10^6$ cells: 0.5 x$10^6$ *Mtbc* bacilli) and the numbers of intracellular bacteria (CFU) were determined immediately after uptake at 4h post-infection. A single dot represents the mean of three CFU measurements per infection well therefore three dots represent three infections well replicates (n=3) per strain and, per donor in panel **a**, **b**, **d** and e while it represents the mean of three infection wells per experiment in figures **c** and **f**. Mean and standard deviation are depicted in the figures. Abbreviations: CFU, Colony Forming Unit; MOI, Multiplicity of Infection.

**Suppl. Table S1**: ***M. tuberculosis* complex lineages and sub-lineages considered geographically restricted.**

| Lineage | Simpsons diversity index |
|---|---|
| 1 | - |
| 2.1 (‘Proto-Beijing’) | 0.19 |
| 2.2.2 (‘Asia Ancestral 1’) | 0.67 |
| 2.2.1.1.2 (‘Asia Ancestral 2’) | 0.28 |
| 3 | - |
| 4.3.i2 | 0.25 |
| 4.11 | 0.11 |
| 4.5 | - |
| 4.6.2.2 (‘Cameroon’) | 0.28 |
| 4.6.1.1.1 (‘Uganda’) | 0.28 |
| 4.2.1.1 | 0.09 |
| 5 (*M. africanum* West African 1) | - |
| 6 (*M. africanum* West African 2) | - |

Simpsons diversity index based on Freschi et al. (2).

**Suppl. Table S2**: **Tuberculosis index case and contact characteristics and their association with transmission using Generalized Estimation Equations** (N = 12,749).

| | Tuberculosis Infection | | | Secondary Active Tuberculosis | | |
|---|---|---|---|---|---|---|
| Characteristic | OR[1] | 95% CI[1] | p-value | OR[1] | 95% CI[1] | p-value |
| Age (contact) | 1.14 | 1.11, 1.17 | **<0.001** | 0.96 | 0.90, 1.01 | 0.13 |
| Sex (contact) | 1.01 | 0.92, 1.10 | 0.9 | 1.71 | 1.44, 2.04 | **<0.001** |
| High-inc. country born (contact) | 2.18 | 1.96, 2.42 | **<0.001** | 0.61 | 0.49, 0.76 | **<0.001** |
| Age (index) | 0.91 | 0.88, 0.93 | **<0.001** | 0.77 | 0.72, 0.83 | **<0.001** |
| Smear positive (index) | 1.48 | 1.30, 1.68 | **<0.001** | 0.94 | 0.70, 1.26 | 0.7 |
| Alcohol use (index) | 1.01 | 0.77, 1.32 | >0.9 | 3.06 | 1.73, 5.42 | **<0.001** |
| Drug use (index) | 0.95 | 0.67, 1.34 | 0.8 | 0.85 | 0.47, 1.54 | 0.6 |
| Geographically restricted lineage (index) | 0.82 | 0.72, 0.94 | **0.004** | 0.77 | 0.57, 1.04 | 0.088 |

[1]OR = Odds Ratio, CI = Confidence Interval, P-values from Wald test

**Suppl. Table S3**: **New York City tuberculosis index case and contact characteristics and their association with transmission using Generalized Estimation Equations** (N = 8,481).

| | Tuberculosis Infection | | | Secondary Active Disease | | |
|---|---|---|---|---|---|---|
| Characteristic | OR[1] | 95% CI[1] | p-value | OR[1] | 95% CI[1] | p-value |
| Age (contact) | 1.12 | 1.09, 1.16 | **<0.001** | 0.84 | 0.78, 0.92 | **<0.001** |
| Sex (contact) | 0.95 | 0.85, 1.05 | 0.3 | 1.67 | 1.29, 2.16 | **<0.001** |
| High-inc. country-born (contact) | 2.24 | 1.97, 2.55 | **<0.001** | 1.93 | 1.39, 2.68 | **<0.001** |
| HIV (contact) | 0.99 | 0.57, 1.70 | >0.9 | 1.28 | 0.41, 3.98 | 0.7 |
| Diabetes (contact) | 1.56 | 1.15, 2.12 | **0.004** | 4.23 | 2.36, 7.59 | **<0.001** |
| Age (index) | 0.91 | 0.88, 0.95 | **<0.001** | 0.95 | 0.87, 1.03 | 0.2 |
| Smear positive (index) | 1.62 | 1.38, 1.90 | **<0.001** | 1.14 | 0.79, 1.66 | 0.5 |
| Alcohol use (index) | 0.73 | 0.48, 1.09 | 0.13 | 2.24 | 1.05, 4.78 | **0.037** |
| Drug use (index) | 0.98 | 0.67, 1.44 | >0.9 | 2.04 | 1.21, 3.44 | **0.007** |
| Geographically restricted lineage (index) | 0.92 | 0.78, 1.08 | 0.3 | 0.54 | 0.37, 0.78 | **0.001** |

[1]OR = Odds Ratio, CI = Confidence Interval, P-values from Wald test

**Suppl. Table S4: Associations of study members' characteristics and bacterial lineage on transmission** (N = 2,279).

| | Tuberculosis Infection | | | Secondary Active Tuberculosis | | |
|---|---|---|---|---|---|---|
| Characteristic | IRR[1] | 95% CI[1] | p-value | IRR[1] | 95% CI[1] | p-value |
| Age (index) | 0.97 | 0.95, 0.99 | **0.003** | 0.90 | 0.81, 1.00 | **0.043** |
| Sex (index) | 1.04 | 0.94, 1.14 | 0.5 | 1.15 | 0.76, 1.78 | 0.5 |
| Smear positive (index) | 1.34 | 1.21, 1.48 | **<0.001** | 1.41 | 0.90, 2.31 | 0.15 |
| Alcohol use (index) | 1.09 | 0.87, 1.35 | 0.4 | 2.21 | 1.11, 4.01 | **0.015** |
| Drug use (index) | 0.91 | 0.72, 1.14 | 0.4 | 1.32 | 0.60, 2.56 | 0.5 |
| High incidence contact group (%) | 2.01 | 1.76, 2.30 | **<0.001** | 0.91 | 0.50, 1.62 | 0.8 |
| One child contact (index) | 1.03 | 0.94, 1.13 | 0.5 | 0.70 | 0.46, 1.05 | 0.092 |
| Mtbc lineage (index) | | | | | | |
| $L4_{widespread}$ | — | — | | — | — | |
| L1 | 0.71 | 0.60, 0.85 | **<0.001** | 0.23 | 0.05, 0.69 | **0.027** |
| $L2_{restricted}$ | 0.87 | 0.62, 1.17 | 0.4 | 1.02 | 0.23, 2.89 | >0.9 |
| $L2_{widespread}$ | 0.96 | 0.85, 1.08 | 0.5 | 0.70 | 0.38, 1.22 | 0.2 |
| L3 | 0.91 | 0.78, 1.05 | 0.2 | 0.41 | 0.16, 0.88 | **0.038** |
| $L4_{restricted}$ | 0.98 | 0.77, 1.22 | 0.8 | 1.27 | 0.49, 2.75 | 0.6 |
| L5/L6 | 0.95 | 0.61, 1.40 | 0.8 | | | |

[1]IRR = Incidence Rate Ratio, Mtbc = M. tuberculosis complex, CI = Confidence Interval, P-values from Wald test

**Suppl. Table S5: Contact type stratified by allopatric and sympatry exposure for contact of tuberculosis index cases in New York City**

| Characteristic | N | **Allopatric exposure** N = 2,495[1] | **Sympatric exposure** N = 5,166[1] |
|---|---|---|---|
| **Contact type** | 7,661 | | |
| Missing | | 1 (<0.1%) | 0 (0%) |
| Health care | | 174 (7.0%) | 266 (5.1%) |
| Homeless | | 0 (0%) | 15 (0.3%) |
| Household | | 1,404 (56%) | 2,971 (58%) |
| Leisure | | 99 (4.0%) | 283 (5.5%) |
| Other | | 29 (1.2%) | 169 (3.3%) |
| School / Daycare | | 253 (10%) | 391 (7.6%) |
| Workplace | | 522 (21%) | 1,070 (21%) |
| Worship | | 13 (0.5%) | 1 (<0.1%) |

[1]Median (IQR); n (%), TB = Tuberculosis

**Suppl. Table S6: Effect of contact allopatry on infection assuming different country-lineage sympatry relationships.**

| | Sympatry for *Mtbc* L1,L3,L5,L6 | | | | plus *Mtb* L2 ancestral (2.1, 2.2.2, 2.2.1.1.2 in East Asia) | | | | plus *Mtb* L4 restricted sublineages (4.3.i2, 4.11, 4.5) | | | | plus *Mtb* L2 modern (in East Asia) | | | |
|---|---|---|---|---|---|---|---|---|---|---|---|---|---|---|---|---|
| Characteristic | N | OR[1] | 95% CI[1] | p-value | N | OR[1] | 95% CI[1] | p-value | N | OR[1] | 95% CI[1] | p-value | N | OR[1] | 95% CI[1] | p-value |
| High incidence birth (contact) | 2,556 | 2.34 | 1.62, 3.39 | **<0.001** | 2,788 | 2.35 | 1.63, 3.38 | **<0.001** | 2,988 | 2.37 | 1.74, 3.24 | **<0.001** | 4,830 | 3.10 | 2.39, 4.02 | **<0.001** |
| Allopatry (contact) | 2,556 | 0.62 | 0.43, 0.91 | **0.013** | 2,788 | 0.66 | 0.46, 0.95 | **0.025** | 2,988 | 0.68 | 0.50, 0.92 | **0.014** | 4,830 | 1.06 | 0.83, 1.36 | 0.6 |
| Age (contact) | 2,556 | 1.14 | 1.07, 1.21 | **<0.001** | 2,788 | 1.15 | 1.08, 1.22 | **<0.001** | 2,988 | 1.14 | 1.07, 1.20 | **<0.001** | 4,830 | 1.13 | 1.08, 1.18 | **<0.001** |
| Sex (contact) | 2,556 | 0.97 | 0.79, 1.18 | 0.8 | 2,788 | 0.95 | 0.79, 1.15 | 0.6 | 2,988 | 0.96 | 0.80, 1.15 | 0.6 | 4,830 | 1.07 | 0.93, 1.22 | 0.4 |
| Smear positivity (index) | 2,556 | 1.78 | 1.35, 2.36 | **<0.001** | 2,788 | 1.80 | 1.37, 2.37 | **<0.001** | 2,988 | 1.60 | 1.24, 2.05 | **<0.001** | 4,830 | 1.49 | 1.22, 1.82 | **<0.001** |
| Alcohol (index) | 2,556 | 0.90 | 0.34, 2.43 | 0.8 | 2,788 | 1.13 | 0.53, 2.41 | 0.7 | 2,988 | 1.02 | 0.50, 2.07 | >0.9 | 4,830 | 0.69 | 0.40, 1.19 | 0.2 |
| Drug use (index) | 2,556 | 1.41 | 0.71, 2.80 | 0.3 | 2,788 | 1.19 | 0.63, 2.25 | 0.6 | 2,988 | 1.31 | 0.72, 2.35 | 0.4 | 4,830 | 1.07 | 0.58, 1.96 | 0.8 |

[1]OR = Odds Ratio, CI = Confidence Interval, *Mtbc* = *M. tuberculosis* complex

**Suppl. Table S7: Generalized estimation equation model to quantify the effect of contact sympatry on *M. tuberculosis* complex infection among New York City contacts exposed to a strain of a geographically restricted lineage** (N = 2,012).

| Characteristic | OR[1] | 95% CI[1] | p-value |
|---|---|---|---|
| Age (contact) | 1.11 | 1.04, 1.19 | **0.002** |
| Sex (contact) | 0.92 | 0.73, 1.16 | 0.5 |
| High-inc. country-born (contact) | 2.27 | 1.56 3.22 | **<0.001** |
| Allopatry (contact) | 0.068 | 0.48, 0.96 | **0.03** |
| HIV (contact) | 0.00 | 0.00, 0.00 | **<0.001** |
| Diabetes (contact) | 1.01 | 0.54, 1.89 | 0.9 |
| Smear positive (index) | 1.75 | 1.31, 2.34 | **<0.001** |
| Alcohol (index) | 0.83 | 0.37, 1.86 | 0.7 |
| Drug use (index) | 1.82 | 0.98, 3.38 | 0.059 |

[1]OR = Odds Ratio, CI = Confidence Interval, P-values from Wald test

**Suppl. Table S8: Generalized estimation equation model to quantify the effect of exposure time on *M. tuberculosis* complex infection among tuberculosis index case contacts with exposure time available** (N = 22,145).

| Characteristic | OR[1] | 95% CI[1] | p-value |
|---|---|---|---|
| Age (contact) | 1.13 | 1.1,1.15 | **<0.001** |
| Sex (contact) | 1.08 | 1,1.16 | **0.047** |
| High-inc. country-born (contact) | 2.24 | 2.06,2.82 | **<0.001** |
| Significant exposure time (>8h) (contact) | 2.41 | 2.06,2.82 | **<0.001** |
| Age (index) | 0.93 | 0.9,0.95 | **<0.001** |
| Sputum microscopy (index | 1.4 | 1.24,1.58 | **<0.001** |
| Alcohol (index) | 1.17 | 0.92,1.47 | 0.2 |
| Drug use (index) | 0.91 | 0.66,1.25 | 0.6 |

[1]OR = Odds Ratio, CI = Confidence Interval, P-values from Wald test

#### References Supplementary material


1. D. Klinkenberg, J. A. Backer, X. Didelot, C. Colijn, J. Wallinga, Simultaneous inference of phylogenetic and transmission trees in infectious disease outbreaks. PLoS Comput. Biol. **13**, e1005495 (2017).

2. L. Freschi, R. Vargas Jr, A. Husain, S. M. M. Kamal, A. Skrahina, S. Tahseen, N. Ismail, A. Barbova, S. Niemann, D. M. Cirillo, A. S. Dean, M. Zignol, M. R. Farhat, Population structure, biogeography and transmissibility of Mycobacterium tuberculosis. Nat. Commun. **12** (2021), doi:10.1038/s41467-021-26248-1.
